# Contusion Volume is a Cross-cohort Predictor of Delayed Seizures after Traumatic Brain Injury

**DOI:** 10.64898/2026.06.01.26354621

**Authors:** Ajitesh Nanda, Xiaoying Sun, Frederic L.W.V.J. Schaper, Jennifer A Kim, Hui Shi, Shira Cohen-Zimerman, Amy J Markowitz, Eric S Rosenthal, Michael D Fox, Brian L Edlow, Jordan Grafman, Geoffrey T Manley, Joseph T Giacino, Sonia Jain, Yelena G Bodien, Samuel Snider, TRACK-TBI Investigators

## Abstract

**Objective:** Predicting specific cognitive, psychiatric, and health-related sequelae in patients after acute traumatic brain injury (TBI) remains an important but challenging clinical problem. Acute phase computed tomography (CT) scans acquired show hemorrhagic contusions, a common type of traumatic pathology. However, whether CT-measured contusions predict long-term sequelae is uncertain.

**Methods:** We established a Screening Cohort of patients with acute TBI who received care at a single TBI Model Systems (TBIMS) inpatient rehabilitation facility. Regions of hemorrhagic contusion and edema were labeled on acute brain CT scans using the fully-automated Brain Lesion Analysis and Segmentation Tool (BLAST-CT). We screened 198 outcome variables at 1-year post-injury for association with acute hemorrhagic contusion volume using the Harrell’s Concordance index (C-index), controlling for multiple comparisons using 5,000 outcome permutations. Finally, we tested whether the significant associations in the TBIMS database replicated in acute (Transforming Research and Clinical Knowledge in TBI [TRACK-TBI]) and chronic (Vietnam Head Injury Study [VHIS]) external validation cohorts.

**Results:** The TBIMS Screening Cohort included 345 participants (mean ± SD age: 55.7 ± 21.5 years) with median [IQR] contusion volume 2.3 cc [0.1, 14.6]. Among 198 candidate outcome variables, only delayed seizures were significantly associated with acute hemorrhagic contusion volume (C-index = 0.81; P_FWE_ = 0.007). Contusion volume was not significantly associated with commonly-used measures of global functioning like the Glasgow Outcome Scale Extended, (C-index = 0.55; P_FWE_ = 1). Within the screening cohort, 30 ccs was the optimal volume threshold for discriminating patients with versus without delayed seizures (OR 12.6, 95% CI: [4.6, 34.3]). Contusions larger than 30 cc remained significantly associated with delayed seizures in two external cohorts: (TRACK-TBI OR 4.1 [1.5, 11.2]; VHIS OR 3.2 [1.7, 6.2]).

**Interpretation:** Across three cohorts of patients with TBI, CT-derived contusion volume is robustly associated with the development of delayed seizures, in contrast to commonly-used outcomes measuring global functioning. A 30-cc volume threshold can be used to improve epilepsy prediction models and enrich populations for clinical trials.

## Introduction

Traumatic brain injury (TBI) is a significant contributor to the global burden of neurological disease, with more than 50 million injuries causing over 8-million person years of disability annually^1,2^. Disability after TBI results from such wide-ranging sequelae as cognitive impairment^3–5^, executive dysfunction^3,4^, impaired strength^6–8^, movement disorders^9–12^, psychiatric disturbances^13–15^, and epilepsy^16–20^. Accurate prediction of specific sequalae in individual patients would enable enhanced monitoring, prophylactic interventions, targeted rehabilitation, and clinical trial enrichment.

Due to the heterogeneity of traumatic brain injuries and their associated sequalae, developing accurate predictive models requires large samples. While MRI is the most sensitive tool for detecting structural brain injuries, building large cohorts of patients imaged acutely with MRI is challenging. In contrast to MRI, CT imaging is fast, widely available, and sensitive for hemorrhagic contusions, the most common type of parenchymal pathology after moderate or severe TBI^21,22^. Nearly every patient diagnosed with moderate or severe TBI has a CT scan acquired as part of routine clinical care. Open-source, artificial intelligence-powered tools now enable fully automated labeling of hemorrhagic contusions on TBI scans. These new tools open the door to investigating the neuroanatomical basis of specific post-TBI sequelae on a previously unattainable scale.

Although automated tools can now identify contusions on acute CT scans, it remains unclear whether contusion size is useful for predicting long-term outcomes. Existing studies are conflicted as to whether CT-derived contusion volume is associated with long-term disability^23,24^, or whether its prognostic utility is limited to acute mortality^25,26^. No studies have yet examined whether CT-derived contusions can predict specific symptoms or sequelae beyond 6 months post-injury.

Here, we analyzed CT scans from more than 850 patients across three of the largest studies of TBI in the United States (US), (TBI Model Systems [TBIMS], Transforming Research and Clinical Knowledge in Traumatic Brain Injury [TRACK-TBI], and the Vietnam Head Injury Study [VHIS]). Our goal was to identify the post-traumatic symptoms or sequelae with the strongest associations to the volume of contusions measured on CT scans, and to quantify the performance of specific predictive thresholds across heterogeneous datasets.

## Methods

### Study Cohorts

We conducted a secondary analysis of data from three large-scale US-based studies of TBI, using one study for screening (TBIMS) and two for validation (TRACK-TBI, VHIS). The Screening Cohort included participants from the Spaulding-Harvard site of the TBIMS Study. TBIMS is a prospective, longitudinal, multi-center study in the US of adult (≥ 16 years) patients with moderate and severe TBI (Mayo Classification^27^) who survive acute care hospitalization and undergo inpatient rehabilitation. Detailed descriptions of the study and its full eligibility criteria have been previously published^28–30^. TRACK-TBI is a prospective multi-center study that enrolls patients with acute TBI at 18 level 1 trauma centers in the US (clinicaltrials.gov: NCT02119182)^31,32^. TRACK-TBI includes a representative sample of patients with acute TBI arriving to acute care hospitals, as opposed to just those who survive and are admitted to an inpatient rehabilitation facility. VHIS is a prospective, longitudinal follow-up study of US Vietnam War veterans who sustained penetrating head trauma during the conflict (clinicaltrials.gov: NCT00132249)^33,34^.

Inclusion criteria for Screening Cohort (TBIMS) subjects for this analysis were: 1) CT scan acquired within 14 days of injury; and 2) outcomes data available 1-year post injury, defined as at minimum having a Glasgow Outcome Scale - Extended (GOSE) assessment performed (Figure 1). Cohort-specific inclusion and exclusion criteria are provided in Supplementary Figures 1 and 2 for TRACK-TBI and VHIS respectively. Overall, 345 participants in the TBIMS Screening Cohort, 390 participants in the TRACK-TBI Validation Cohort, and 161 participants in the VHIS Validation Cohort were included in the final analysis.

**Figure 1.**
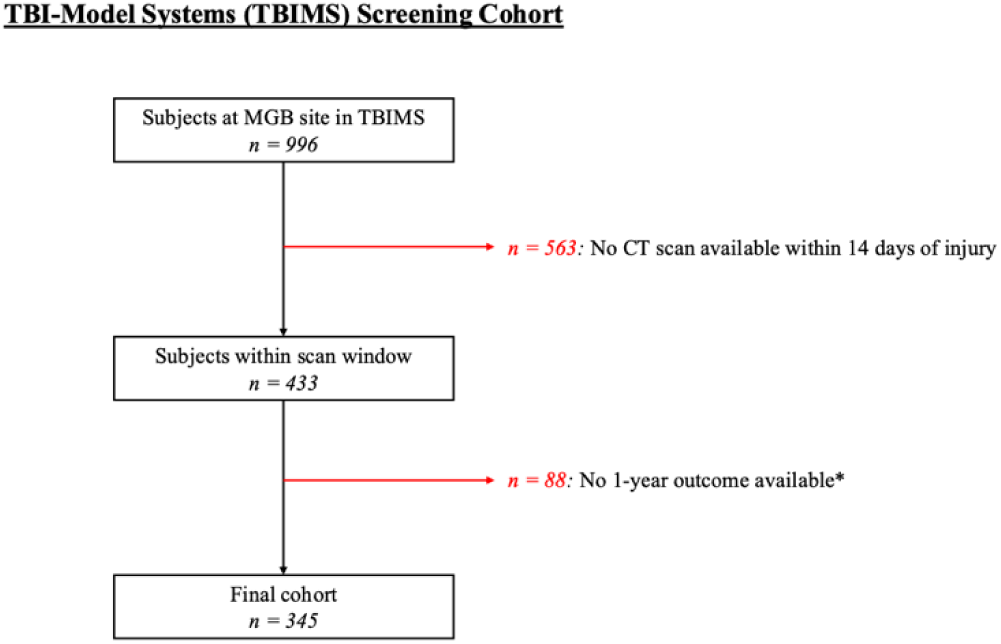
Screening Cohort Consort Diagram. Participants were excluded if a GOSE assessment was not completed at the 1-year study visit. Abbreviations: MGB Mass General Brigham

TBIMS was selected as the Screening Cohort due to the comprehensiveness of its outcome battery and its requirement that participants are admitted to inpatient rehabilitation, minimizing bias from censoring due to early mortality. Significant findings from the TBIMS Screening Cohort were validated in two independent datasets, intentionally selected based on differences with TBIMS. The specific goal was to determine whether findings replicated in TRACK-TBI, a representative population of all TBI patients treated at Level 1 academic acute care hospitals, and VHIS, a sample of patients with chronic, penetrating TBI and longitudinal follow-up for 35 years post-injury.

### CT Processing and hemorrhagic contusion labeling

For TBIMS and TRACK-TBI, we analyzed 5mm-thickness axial slices from non-contrast CT brain scans. If no 5mm acquisition was available, we analyzed 2mm- or 3mm-thickness slices. Because contusions and edema often expand from their initial appearance, we analyzed the latest scan within 48 hours of injury ^35,36^. If no scan was available within 48 hours of injury, we analyzed the earliest scan available within two weeks of injury in TBIMS or three weeks of injury in TRACK-TBI. Raw CT images from TBIMS and TRACK-TBI were processed using BLAST-CT. BLAST-CT is a neural network trained on TBI CT scans that automatically segments four types of traumatic pathology (Figure 2A): 1) intraparenchymal hemorrhage, 2) extra-axial hemorrhage, 3) perilesional edema, and 4) intraventricular hemorrhage (https://github.com/biomedia-mira/blast-ct/)^37^. Because contusions contain intermingled hyperdense blood products and hypodense edema^38,39^, we defined contusion volume as previously^23^: the combined volume of intraparenchymal hemorrhage and edema (Figure 3A).

**Figure 2.**
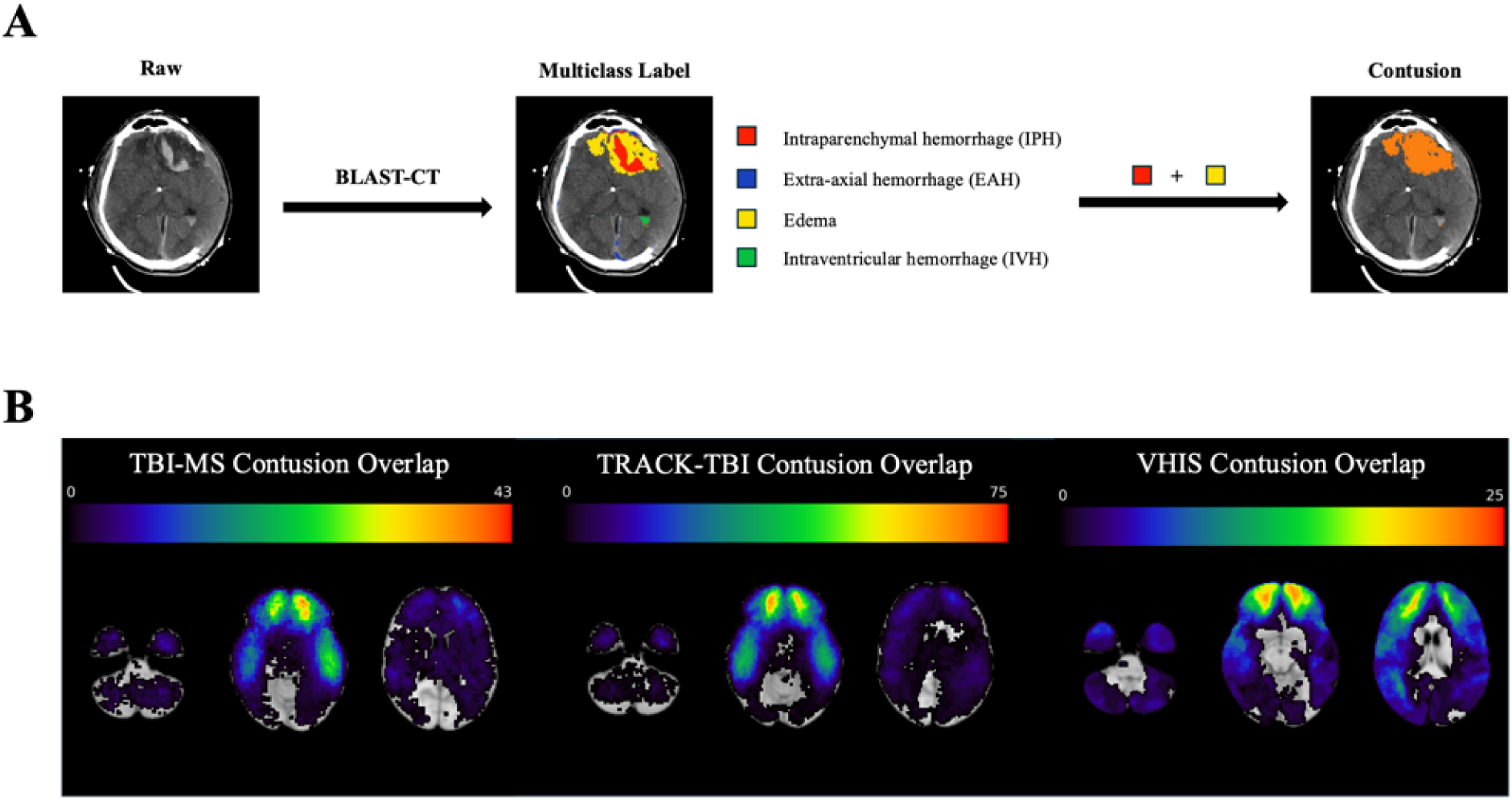
Contusion labeling. **A)** Raw CT scans were processed with BLAST-CT in TBIMS and TRACK-TBI to obtain multiclass lesion labels. IPH and edema labels were combined into an overall contusion measurement for analysis. In VHIS, encephalomalacic lesions were manually traced. All scans were registered into MNI T1 2mm space. **B)** MNI-space contusion overlap maps are shown for each cohort, where the color represents number of participants with a contusion at each voxel.

**Figure 3.**
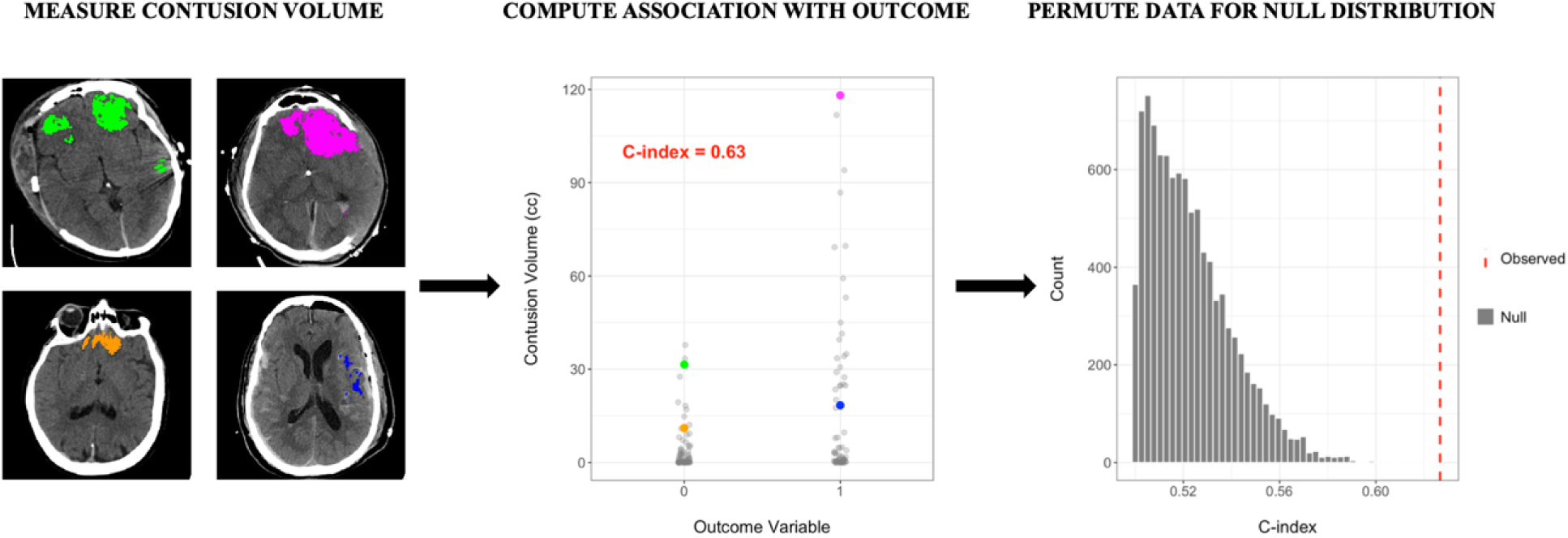
Methodology of outcome association screen. For each subject, contusion volumes are measured with BLAST-CT and mapped to their response for a given outcome variable, yielding an observed C-index. Using randomly permuted outcome values, this procedure is repeated 5,000 times across all outcome variables, yielding a null distribution of the most-extreme C-indices. The observed C-index is then compared to this null distribution to assign a p-value.

For VHIS, 2.5 millimeter-thickness axial CT scans were obtained as part of the Phase 3 follow-up assessment, 35 years after the injury. Encephalomalacic lesions resulting from the initial TBI were manually traced on the CT scans by a trained neuropsychiatrist and validated by an independent, blinded researcher, as described previously^40^.

All CT scans were registered to Montreal Neurological Institute Space T1 2mm space for visualization purposes (Supplementary Methods: Figure 2B).

### Outcomes

TBIMS collects annual comprehensive outcomes data following the initial injury. These outcomes include variables that assess global level of independence, cognition, behavioral function, motor function, psychiatric symptoms, medical and neurological disorders, and quality of life metrics. For this analysis, all outcome data was compiled from the 1-year follow up visit. From 315 variables collected in TBIMS, we identified the subset of 198 with potential to be a sequela of parenchymal injury, whose responses were binary, ordinal, or continuous, and for which there were at least 50 non-missing values to analyze. For binary or ordinal variables, we required at least 5 observations in the nondominant class to ensure sufficient variance. Where relevant, responses were aggregated across multiple variables to create a single summary variable. A detailed description of all screened variables in TBIMS is provided in the Supplementary Methods.

For validation in TRACK-TBI, we analyzed a specific outcome available at either 6-months or 1-year post injury. For validation in VHIS, we analyzed the incidence of this outcome at 35 years post injury. A detailed description of the outcomes analyzed in each cohort is provided in the Supplementary Methods.

### Statistical analysis: Lesion Volume and Outcomes

In the Screening Cohort (TBIMS), we conducted a screen to measure the association between acute contusion volume and each outcome (Figure 3). The primary measure of association was Harrell’s concordance index (C-index), which quantifies whether the ranking of a pair of subjects by the dependent variable is concordant with the ranking by the independent variable^41^. The C-index provides a generalizable form of the area under the receiving operating characteristic (AUC) for continuous, ordinal, and binary variables^42^. To define a study-level null distribution with control of the family-wise error (FWE) rate, we performed 5,000 permutations of all outcome variables. From each permutation, the most extreme C-index across all outcomes was recorded and used to build the final null distribution. Empirical p-values were then assigned to each outcome by comparing each’s true C-index to this global null distribution, with significance set at P_FWE_ < 0.05. For significant predictors of binary outcomes, we identified the Youden-optimal predictive threshold and reported the performance characteristics. Because the C-index may be differentially sensitive to continuous versus binary outcomes, we repeated the screen using linear or logistic regression as appropriate and compared the test statistic for each predictor’s regression coefficient (Supplementary Methods: Regression Screen).

### External Validation

We report crude odds ratios (OR) for contusion volume in TRACK-TBI and VHIS for any significant outcomes from the TBIMS screen, as well as the performance characteristics of the TBIMS-identified optimal volume cut-points.

All statistical analyses were performed in R Studio (Version 2024.12.1+563).

## Results

### Cohort Characteristics

**Table 1.**

| Table 1 |  |
| --- | --- |
|  | TBI-Model Systems (Screening Cohort; N= 345) |
| Age (years)<br>Mean ± SD<br>Total | 55.7 ± 21.5<br>340 |
| Sex<br>Male<br>Female<br>Total | 239 (69.3%)<br>106 (30.7%)<br>345 |
| Race<br>White<br>Black<br>Other<br>Total | 267 (87.0%)<br>21 (6.8%)<br>19 (6.2%)<br>307 |
| Ethnicity<br>Hispanic<br>Non-Hispanic<br>Total | 22 (7.1%)<br>290 (92.9%)<br>312 |
| Education (years)<br>Median [IQR]<br>Total | 13 [12, 16]<br>333 |
| Pre-injury Employment<br>Employed<br>Student<br>Retired | 154 (45.4%)<br>15 (4.4%)<br>134 (39.5%) |
| Unemployed | 17 (5.0%) |
| Other | 19 (5.6%) |
| Total | 339 |
| Marital Status |  |
| Single | 120 (35.2%) |
| Married | 139 (40.8%) |
| Other | 82 (24.0%) |
| Total | 341 |
| Pre-injury Psychiatric History |  |
| Yes | 101 (30.2%) |
| No | 233 (69.8%) |
| Total | 334 |
| Injury Cause |  |
| Motor Vehicle Related | 82 (23.9%) |
| Fall | 189 (55.1%) |
| Assault / Violence | 18 (5.2%) |
| Other | 54 (15.7%) |
| Total | 343 |
| GCS on ED Arrival |  |
| 13-15 | 159 (46.5%) |
| 9-12 | 27 (7.9%) |
| 3-8 | 156 (45.6%)* |
| Total | 342 |
| Median [IQR] | 11 [3,14] |
| SAH |  |
| Yes | 266 (78.0%) |
| No | 75 (22.0%) |
| Total | 341 |
| Contusion Volume (cc) |  |
| Median [IQR] | 2.28 [0.15, 14.58] |
| Total | 345 |
| GOSE at 1-year |  |
| 5-8 | 204 (59.1%) |
| 2-4 | 107 (31.0%) |
| 1 | 34 (9.9%) |
| Total | 345 |
| * In TBIMS, participants who were chemically paralyzed (n = 56) or intubated (n = 68) did not receive a GCS score. They were assigned a score of GCS 3 for reporting in Table 1. |  |

The Screening Cohort (TBIMS) included N=345 participants with TBI enrolled at a single academic medical center between 2013 and 2023 with a CT scan acquired during the acute care hospitalization. The mean (± SD) age was 55.7 (± 21.5) years, 69% were male and 87% were white. The most frequent mechanism of injury (55%) was fall. The Glasgow Coma Scale (GCS) score on admission to acute care hospital was 13-15 in 47%, 9-12 in 8%, and <9 in 46% of participants. The median BLAST-CT measured contusion volume was 2.3 cc, IQR [0.1, 14.6] and contusions tended to occur in the inferior frontal and temporal lobes (Figure 2B).

### Association between contusion volume and 1-year outcomes

Among 198 candidate outcome variables, the only outcome significantly associated with contusion volume after controlling for multiple comparisons was the occurrence of any seizures between 7 days and 1 year post-injury (C-index = 0.81, P_FWE_ = 0.007, n = 238; Figure 4). The outcomes with the next-strongest association to contusion volumes, though not significant after controlling for multiple comparisons, included four items on the Disability Rating Scale measuring communication ability and cognition required for self-care (C-indices = 0.70-0.72; P_FWE_ = 0.15-0.26; n = 303-310). Contusion volume was not significantly associated with the GOSE, the most common measure of global functioning after TBI, (C-index = 0.55; P_FWE_ = 1; n = 345). Results were consistent when repeating the screen using regression models instead of C-indices (Supplementary Methods: Supplementary Figure 3).

**Figure 4.**
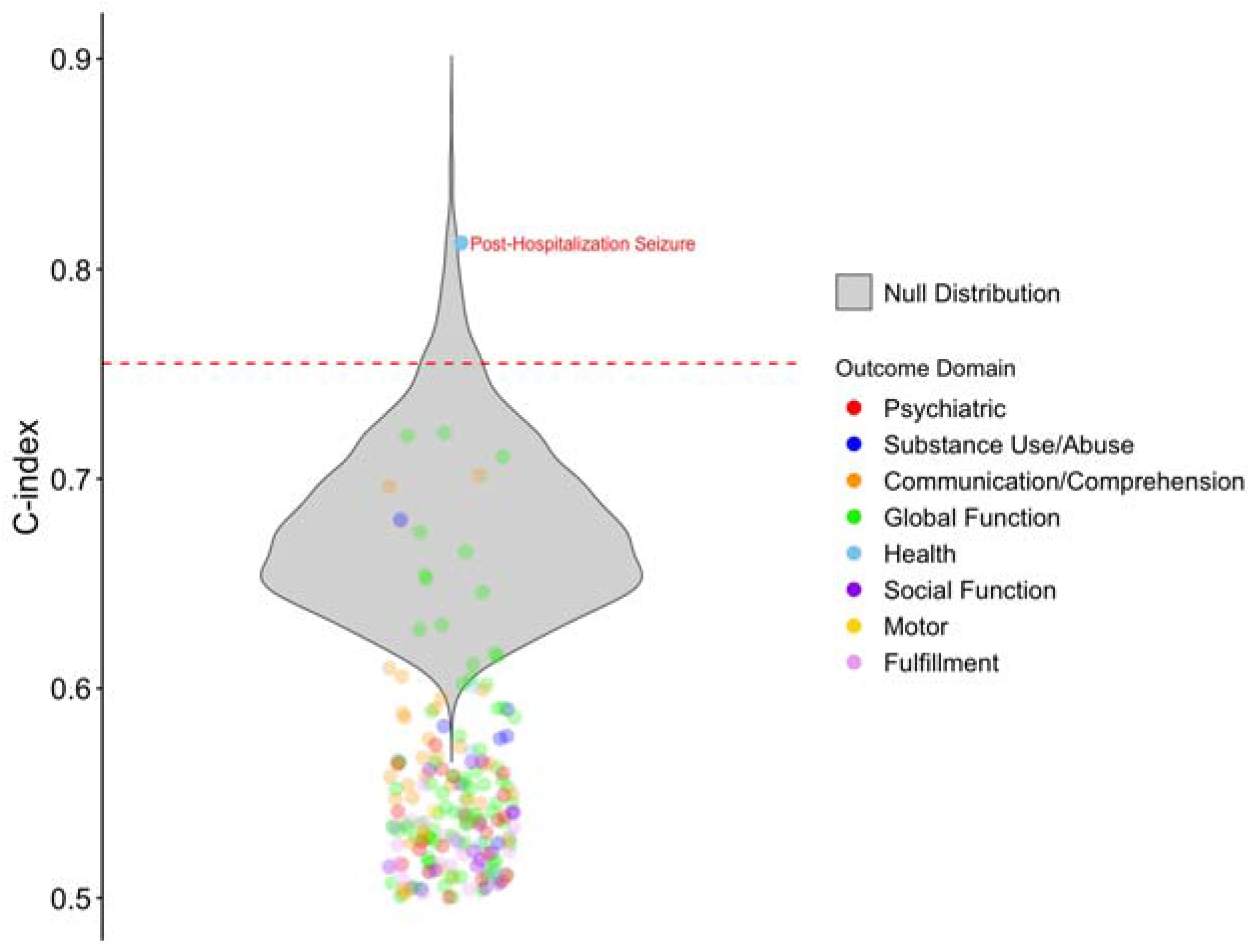
**Screening Cohort outcome association results**The observed C-indices are plotted for each outcome variable over the null distribution of 5,000 most extreme C-indices across all permutations. Outcomes are grouped and colored by study-defined domains. The red line represents the family-wise error threshold for significance (P < 0.05). C-indices indicating negative association (C-index < 0.5) have been flipped to 1 – C-index such that all C-indices are reported as greater than 0.5, analogous to AUC.

The Youden optimal threshold contusion volume for predicting delayed seizures was 30 ccs, where the accuracy was 85%, sensitivity was 0.65, specificity was 0.87, positive predictive value (PPV) was 0.32 and negative predictive value (NPV) was 0.97. Among participants with contusions ≤ 30 ccs, 3% developed seizures, while among participants with > 30 cc contusions, 32% developed seizures (odds ratio [OR] 12.6, 95% CI [4.6, 34.3]; p < 0.0001). Neither age (p = 0.34) nor admission GCS (p = 0.11) differed among participants who did or did not develop seizures (Supplementary Table 1). Patients who developed seizures were more likely to have had a seizure during hospitalization (p = 0.02; Supplementary Table 1). Results were consistent after excluding participants with documented (n=9) or missing data (n=62) on pre-injury seizure disorders (Supplementary Table 2), and extending follow-up to 5-years post injury (Supplementary Table 2).

### External validation of contusion volume threshold

We quantified the performance of the 30 cc contusion volume threhsold for predicting delayed post-traumatic seizures in two external datasets. Compared to TBIMS, TRACK-TBI participants (n=390; Supplementary Table 3) tended to be younger, have lower presenting GCS scores, CT scans acquired earlier post injury, and smaller, though similarly-distributed contusions (Figure 2B). The presence of contusions larger than 30 ccs remained a significant predictor of developing seizures 7-days to 1 year post injury with 88% accuracy (OR 4.1; 95% CI [1.5, 11.2]; p = 0.006; sensitivity = 0.16, specificity = 0.95, PPV = 0.27, NPV = 0.92). 8% of participants with ≤ 30cc contusions developed seizures by 12 months, compared with 27% of those with contusions > 30 cc (Figure 5B). Results were consistent when restricting outcome assessments to the first 6-months post injury (Supplementary Table 4). Neither age (p = 0.52) nor admission GCS (p = 0.56) differed among participants who did or did not develop seizures in TRACK-TBI (Supplementary Table 5).

**Figure 5.**
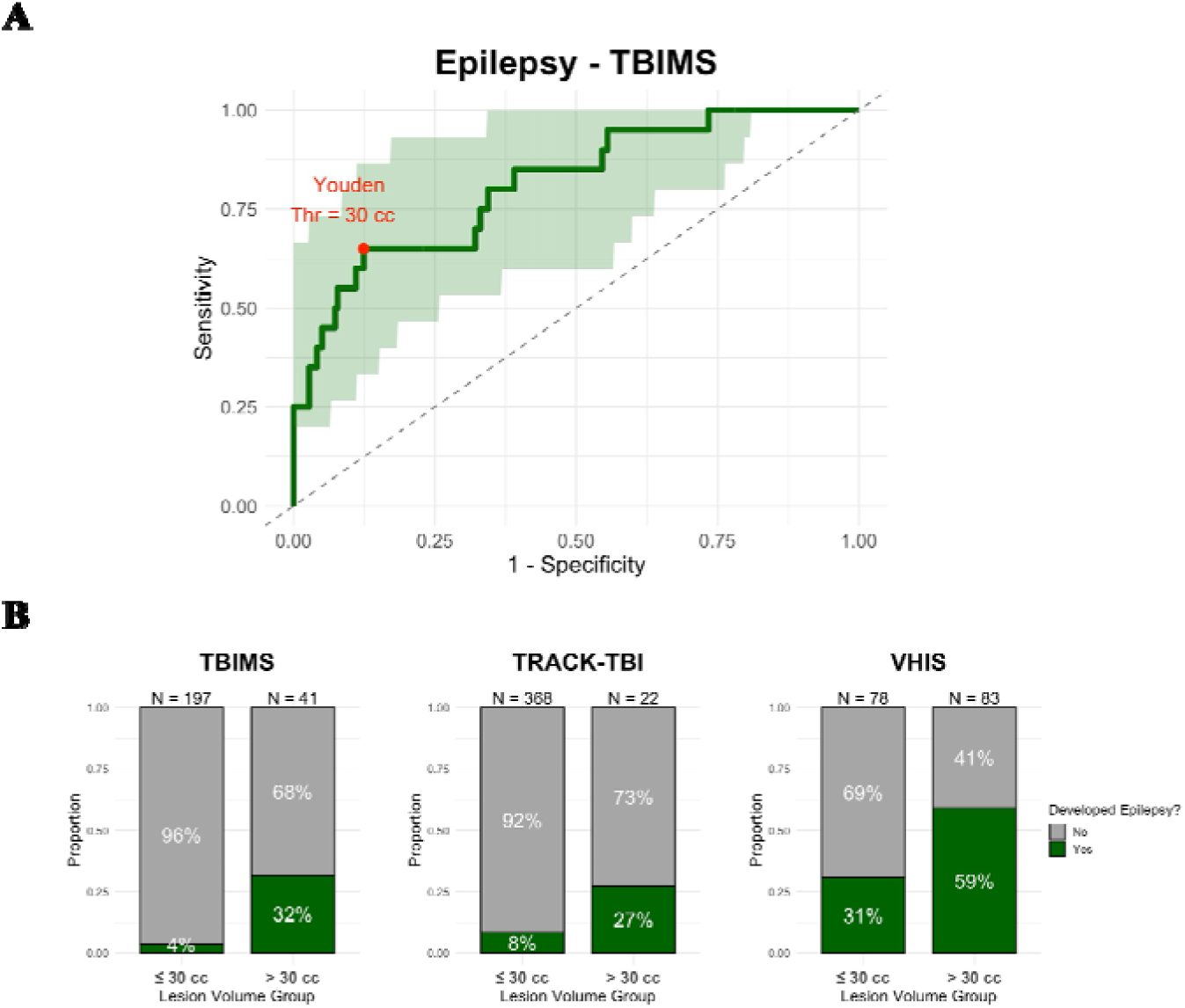
Contusions larger than 30 ccs predict epilepsy across datasets. **A)** An ROC curve was generated using lesion volume to predict epilepsy in the Screening Cohort (AUC = 0.79). The Youden Threshold at 30 cc is annotated on the curve. **B)** For each cohort, subjects have been grouped by lesion volume compared to the threshold with color representing each outcome.

Compared to TBIMS, VHIS participants (n=161, Supplementary Table 6) were younger, all male, and had significantly larger, though similarly-distributed, lesions (Figure 2B). Lesions larger than 30 cc were significantly associated with delayed post-traumatic seizures (OR 3.2; 95% CI [1.7, 6.2]; p = 0.0004; accuracy = 0.64, sensitivity = 0.61, specificity = 0.61, PPV = 0.40, NPV = 0.78). In VHIS, 31% of participants with ≤ 30 cc lesions developed seizures, compared to 59% of those with lesions > 30 cc (Figure 5B). Neither age (p = 0.10) nor duration of loss of consciousness after initial injury (p = 0.51) differed among participants who did or did not develop seizures (Supplementary Table 7).

To assess the reliability of the 30-cc contusion volume threshold across datasets, we measured the incidence of delayed seizures in each dataset in 10 cc buckets of increasing contusion volume. In all 3 datasets, the biggest increase in seizure incidence occurred in the range of 20-30 ccs (Supplementary Figure 4).

## Discussion

Across nearly 200 cognitive, neuropsychiatric, and behavioral outcomes assessed 1-year after TBI, the volume of hemorrhagic contusions on acute CT scans was most associated with delayed seizures. In particular, contusions larger than 30 ccs were robustly associated with the development of delayed post-traumatic seizures in three independent studies, despite each having different approaches to measuring this outcome. Including patients surviving to acute rehabilitation (TBIMS), civilian patients presenting to acute trauma centers (TRACK-TBI), and military patients surviving penetrating injuries (VHIS), these studies represent much of the heterogeneity of TBI. By defining and validating a specific, actionable threshold to predict post-traumatic epilepsy, our findings have implications for the future research and clinical care of patients with TBI.

While existing literature on contusions and outcomes after TBI has focused on global measures of disability^23,24,43^, we instead found the strongest association with delayed post-traumatic seizures. In the acute cohorts, 27-32% of patients with contusions larger than 30 cc developed seizures, compared to 4-8% of those with smaller contusions. The persistence of the association between large contusions and post-acute seizures across extremely heterogenous patient cohorts speaks to the robustness and validity of this finding.

Patients with at least one delayed (> 7 days from injury) post-traumatic seizure are generally considered to have post traumatic epilepsy (PTE) due to their high risk of recurrence, however definitions vary in their requirements for observed recurrence and timing^44–46^. PTE is a well-described complication of TBI, with an estimated incidence of 4% - 25%^16,19,47^ depending on the severity of injury and post-injury assessment window. PTE can substantially impact morbidity and mortality, as well as outcomes of mood, cognition, degree of functional independence and quality of life^48–50^.

Reported risk factors for PTE include severity of TBI^51^, dural penetration^52^, and presence of contusion or subdural hematoma^18^. Early work developing PTE prediction models have incorporated some measure of intracranial injury severity^53–56^.

Associations between acute contusion volume (or surrogate measures) and PTE and have been reported in small cohorts^53,57^, but no study has been able to identify a specific volume threshold value that predicts PTE in independent data. As contusions are common, the lack of a specific, validated volume threshold for predicting PTE is a key barrier to the clinical translation of single sample associations. Our findings extend the existing literature by defining a specific threshold (approximately 30 ccs) that robustly increases PTE risk regardless of type and chronicity of TBI.

The reduced sensitivity of the 30 cc threshold from TBIMS to TRACK-TBI may have been due to systematic differences in how the outcome was assessed, earlier scan times, or a greater prevalence of unmeasured, non-contusive injury. In VHIS, specificity of the 30 cc threshold was reduced relative to TBIMS, likely due to the greater prevalence of PTE in the cohort^58^. The increased prevalence in VHIS may have been due to the penetrating nature of the head trauma, an established risk factor for PTE^52^.

After an acute TBI, current guidelines recommend at most a limited 7-day course use of prophylactic anti-epileptic medication, given the increased risks of adverse effects relative to benefits^59–62^. Our findings may call for a reappraisal of the risk benefit calculations in certain patient subgroups. Relative to those with smaller contusions, patients with large contusions may have a greater than 20% increase in the absolute risk of PTE. In other contexts, an absolute risk difference of this magnitude has been enough to motivate a change in practice. For example, prophylactic anti-epileptic medications are administered following supratentorial but not infratentorial cranial surgery, based on an estimated seizure risk difference of approximately 10% (4% vs 13%)^63^. While previous randomized controlled trials of prophylactic anti-epileptic drugs for preventing early and late seizures demonstrated mixed results^64–66^, these trials used prior-generation anti-epileptics, had poor protocol adherence and follow-up, and had low incidence of delayed seizures^67^. Repeating such trials in populations enriched for risk of delayed seizures with newer anti-epileptic medications should be considered.

Although no specific pathophysiology has linked contusion volume to PTE, the edema and hemorrhage that comprise contusions contain iron compounds (e.g. hemosiderin) and inflammatory markers (e.g., interleukin 1β) which have demonstrated epileptogenicty^68,69^. In animal models, increased traumatic lesion size has been previously demonstrated to be associated with increased seizures^70,71^. Processes such as neurogenesis and excitatory axon sprouting may disrupt regular rhythmicity and synchronization of brain activity^72,73^. The 30-cc threshold may represent a critical point under a multi-hit framework in which the normal balance of excitatory and inhibitory connections tends to become disrupted, increasing susceptibility to overexcitation and seizure. Future studies in TBI should explore a potentially causal relationship between contusion volume, connectivity alterations, and development of PTE.

Finally, in our TBIMS screen, we observed a striking lack of association between contusion volume and several commonly-used global outcome assessment measures, most prominently the GOSE^74,75^. This stands in contrast to existing literature reporting associations between contusion volume and 6-month global outcome measures^23,24,43^. The contrast between previously positive and our negative findings illustrate the outsized role that early mortality may play in driving such associations. In TBIMS, patients who die during the acute hospitalization—typically due to withdrawal of life sustaining therapy—are not enrolled^76,77^. It is possible that the observation of large contusions on early CT may influence clinicians’ prognostic viewpoints and increase likelihood of withdrawal of life sustaining therapy^78^. However, there may also bias in the other direction. By only enrolling participants who attend acute rehabilitation, TBIMS pre-selects patients who are expected to functionally improve, and who will receive a dedicated recovery intervention. In such a population, the influence of contusion volume may be less than among all patients with moderate or severe TBI.

### Limitations

Several important limitations should be acknowledged. First, CT scans are known to be insensitive to certain types of traumatic pathology, particularly diffuse axonal injury. Associations with diffuse axonal injury severity would not be detectable with our screening approach based solely on CT-derived contusion volumes. In VHIS, encephalomalacic lesions were traced if thought to be related to the TBI, including blunt and penetrating injury or surgical debridement, representing a broader set of pathologies than evaluated in TBIMS. Second, our relatively stringent statistical threshold (family wise error < 5%), optimized to identify the strongest and most robust associations, rendered us susceptible to Type II errors. Third, we did not investigate or otherwise correct for other health-modifying events that may have occurred between the initial injury and the data collected at follow-up. For example, participants may have undergone neurosurgery, received other types of critical care interventions, sustained secondary TBI or other injury, or been unable to access adequate follow-up care, each of which could have impacted outcomes independently from the initial TBI. Finally, the delayed post-traumatic seizure assessments differed by study and were reported by patients and/or their caregivers, with only TRACK-TBI requiring a concurrent medical diagnosis. Self-reported outcomes can create bias due to underestimation or overestimation of one’s own ability, social desirability to appear healthier, and recency bias in recalling events over the 1-year period. In the context of PTE, certain unwitnessed seizure types could have gone unrecognized or been mischaracterized.

### Conclusions

CT-derived contusion volume was robustly associated with delayed post-traumatic seizures across three different TBI cohorts. Patients with contusions larger than 30 ccs appear at the highest risk. Based on these data, the 30-cc contusion threshold could be incorporated into individualized PTE prediction models and used for enrichment of clinical trials.

## Data Availability

All data produced in the present study are available upon reasonable request to the principal investigators of the TBIMS, TRACK-TBI, and VHIS studies.

## The TRACK-TBI Investigators

Edilberto Amorim, MD, University of California, San Francisco; Gretchen Brophy, PharmD, Virginia Commonwealth University; Ann-Christine Duhaime, MD, MassGeneral Hospital for Children; Shawn Eagle, PhD, University of Pittsburgh; Brandon Foreman, MD, University of Cincinnati; Ramesh Grandhi, MD MS, University of Utah; Vijay Krishnamoorthy, MD, Duke University; Christine Mac Donald, PhD, University Washington; Debbie Madhok, MD, University of California, San Francisco; Michael McCrea, PhD, Medical College of Wisconsin; Randall Merchant, PhD, Virginia Commonwealth University; Laura B. Ngwenya, MD, PhD, University of Cincinnati; David Okonkwo, MD PhD, University of Pittsburgh; Claudia Robertson, MD, Baylor College of Medicine; Richard B Rodgers, MD, Goodman Campbell Brain and Spine; David Schnyer, PhD, University of Rhode Island; Sabrina R. Taylor, PhD, University of California, San Francisco; Nancy Temkin, PhD, University of Washington; John K. Yue, MD, University of California, San Francisco; Ross Zafonte, DO, University of Missouri

## Acknowledgements

We acknowledge the patients, families, and caregivers who made this work possible.

