## Supplement for "Contusion Volume is a Cross-cohort Predictor of Delayed Seizures after Traumatic Brain Injury"

**Supplementary Material**

**Affiliations**

**1** Center for Brain Circuit Therapeutics, Departments of Neurology, Psychiatry, and Radiology, Brigham and Women’s Hospital, Boston, MA

**2** Biostatistics Research Center, Herbert Wertheim School of Public Health, University of California, San Diego, San Diego, CA

**3** Department of Neurology, Yale University School of Medicine, New Haven, CT

**4** Cognitive Neuroscience Laboratory, Brain Injury Research, Shirley Ryan Ability Lab, Chicago, IL

**5** Department of Physical Medicine and Rehabilitation, Feinberg School of Medicine, Northwestern University, Chicago, IL

**6** Department of Neurological Surgery, University of California, San Francisco, San Francisco, CA

**7** Harvard Medical School, Boston, MA

**8** Center for Neurotechnology and Neurorecovery, Massachusetts General Hospital, Boston, MA

**9** Department of Neurology, Mass General Brigham and Harvard Medical School, Boston, MA

**10** Athinoula A. Martinos Center for Biomedical Imaging, Department of Radiology, Massachusetts General Hospital and Harvard Medical School, Charlestown, MA

**11** Brain and Spinal Cord Injury Center, Zuckerberg San Francisco General Hospital and Trauma Center, San Francisco, CA

**12** Department of Physical Medicine and Rehabilitation, Spaulding Rehabilitation Hospital, Charlestown, MA

**13** Departments of Surgery, Neurological Surgery, and Physical Medicine and Rehabilitation, Vanderbilt University Medical Center, Nashville, TN

**Corresponding Author:**

Samuel B. Snider, MD,

Brigham and Women’s Hospital,

60 Fenwood Rd, Boston, MA 02115

**The TRACK-TBI Investigators**:

Edilberto Amorim, MD, University of California, San Francisco; Gretchen Brophy, PharmD, Virginia Commonwealth University; Ann-Christine Duhaime, MD, MassGeneral Hospital for Children; Shawn Eagle, PhD, University of Pittsburgh; Brandon Foreman, MD, University of Cincinnati; Ramesh Grandhi, MD MS, University of Utah; Vijay Krishnamoorthy, MD, Duke University; Christine Mac Donald, PhD, University Washington; Debbie Madhok, MD, University of California, San Francisco; Michael McCrea, PhD, Medical College of Wisconsin; Randall Merchant, PhD, Virginia Commonwealth University; Laura B. Ngwenya, MD, PhD, University of Cincinnati; David Okonkwo, MD PhD, University of Pittsburgh; Claudia Robertson, MD, Baylor College of Medicine; Richard B Rodgers, MD, Goodman Campbell Brain and Spine; David Schnyer, PhD, University of Rhode Island; Sabrina R. Taylor, PhD, University of California, San Francisco; Nancy Temkin, PhD, University of Washington; John K. Yue, MD, University of California, San Francisco; Ross Zafonte, DO, University of Missouri

**Table of Contents**

Page 3: Supplementary Methods

Page 6: Supplementary Tables

Page 11: Supplementary Figures

Page 14: Supplementary References

Page 15: TBIMS Variable Appendix: 198 TBIMS outcome variable definitions

**Supplementary Methods**

*CT Scan Registration*

CT scans from TBIMS and TRACK-TBI were first registered to a 3mm standard CT template space, and then to the Montreal Neurological Institute T1 2mm reference brain, both steps via a composite 12-parameter affine registration and diffeomorphic warp (antsRegistrationSyNQuick). VHIS CT scans were directly spatially registered to the Montreal Neurological Institute 2mm brain with an automated image registration and 12-parameter affine fit, as previously described^1^.

*TBIMS Epilepsy Outcome*

The epilepsy outcome asked subjects to self-report the number of seizures over the past 1 year since acute care hospital discharge, and sorted responses into ordinal bins of seizure frequency. For analysis, the responses were converted into a binary measure of whether or not any seizure was observed. The original response bins were:

- 0 - no seizures
- 1 - up to three seizures
- 2 - 4-12 seizures
- 3 - at least one seizure monthly
- 4 - at least one seizure weekly
- 5 - at least one seizure daily

*TRACK-TBI Epilepsy Outcome*

Epilepsy in TRACK-TBI was determined by self-reported incidence of seizure from a questionnaire at 6-months or 1-year post injury. Participants were asked to report whether they had experienced symptoms of a seizure (e.g., uncontrolled movements, spacing out, convulsions) or had been told they have a seizure disorder/epilepsy. The seizure(s) must have occurred at least 7 days after the traumatic brain injury and be accompanied by a diagnosis of epilepsy, seizure disorder, or single seizure. Participants with seizures prior to the incident TBI were excluded.

*VHIS Epilepsy Outcome*

Epilepsy in VHIS was determined by self-reported symptoms of seizure during the 35-year period after the principal head injury. Participants with any history of seizures or similar episodes prior to their traumatic brain injury, or with missing data regarding this, were excluded.

*TBIMS Regression Screen*

The 198 outcomes from the C-index screen were also assessed through a regression screen. A logistic regression was fit for variables with two response classes, and a linear regression was fit for variables with more than two responses. Contusion volumes were log-scaled for all regressions, and outcomes in linear regressions were scaled to have mean 0 and standard deviation of 1 to allow for comparable effect sizes. A global null distribution was constructed through 5000 permutations, extracting the largest T statistic observed in any variable from each iteration. Family-wise error significance was set to 0.05.

**Supplementary Tables**

| Supplementary Table 1: TBIMS Epilepsy Cohort Demographics | | | |
| --- | --- | --- | --- |
|  | **No Epilepsy by 1-year**  **(N = 218)** | **Epilepsy by 1-year**  **(N = 20)** | p-value |
| Age (years)  Mean ± SD  Total | 54.2 ± 21.0  215 | 48.6 ± 25.2  20 | 0.2825 |
| Sex  Male  Female  Total | 150 (69.1%)  67 (30.9%)  217 | 19 (95.0%)  1 (5.0%)  20 | 0.0144 |
| GCS on ED Arrival  13-15  9-12  3-8  Total | 94 (43.1%)  17 (7.8%)  107 (49.1%)*  218 | 4 (20.0%)  3 (15.0%)  13 (65.0%)  20 | 0.1092 |
| Seizure during Hospitalization  Yes  No  Total | 24 (14.6%)  140 (85.4%)  164 | 6 (37.5%)  10 (62.5%)  16 | 0.0191 |
| * In TBIMS, participants who were chemically paralyzed did not receive a GCS score. They were assigned a score of GCS 3 for reporting. Chi-squared p-values are reported for categorical variables. | | | |

| Supplementary Table 2: 30cc Threshold Performance in TBIMS | | | |
| --- | --- | --- | --- |
|  | TBIMS  (1 year)  N = 238 | TBIMS  (1 year, excluding seizure history)  N = 167 | TBIMS  (5 year, excluding seizure history)  N = 167 |
| AUC | 0.81 | 0.79 | 0.82 |
| Sensitivity | 0.65 | 0.67 | 0.67 |
| Specificity | 0.87 | 0.85 | 0.86 |
| Odds Ratio for contusions > 30 ccs [95% CI] | 12.6 [4.6, 34.3] | 11.5 [3.2, 41.3] | 12.5 [3.9, 40.2] |
| p value | < 0.0001 | 0.0002 | < 0.0001 |

| Supplementary Table 3: TBIMS and TRACK-TBI Cohort Demographics | | | |
| --- | --- | --- | --- |
|  | **TBI-Model Systems (Screening Cohort; N= 167)** | **TRACK-TBI**  **(External Validation Cohort; N=390)** | p-value |
| Age (years)  Mean ± SD  Total | 53.9 ± 22.4  166 | 37.8 ± 16.0  390 | < 0.0001 |
| Sex  Male  Female  Total | 113 (67.7%)  54 (32.3%)  167 | 297 (76.3%)  93 (23.7%)  390 | 0.0373 |
| Race  White  Black  Other  Total | 133 (82.6%)  10 (6.2%)  18 (11.1%)  161 | 312 (80.4%)  53 (13.7%)  23 (5.9%)  388 | 0.0076 |
| Ethnicity  Hispanic  Non-Hispanic  Total | 13 (7.8%)  154 (92.2%)  167 | 89 (22.9%)  300 (77.1%)  389 | <0.0001 |
| Pre-injury Employment  Employed  Student  Retired  Unemployed  Other  Total | 88 (53.3%)  6 (3.6%)  59 (35.8%)  8 (4.8%)  4 (2.4%)  165 | 270 (73.0%)  22 (6.0%)  22 (6.0%)  56 (15.1%)  0 (0%)  385 | < 0.0001 |
| Pre-injury Psychiatric History  Yes  No  Total | 61 (36.5%)  106 (63.5%)  167 | 84 (21.5%)  306 (78.5%)  390 | 0.0002 |
| Injury Cause  Motor Vehicle Related  Fall  Assault / Violence  Other  Total | 64 (38.3%)  84 (50.3%)  9 (5.4%)  10 (6.0%)  167 | 242 (62.4%)  85 (22.0%)  28 (7.2%)  33 (8.5%)  388 | < 0.0001 |
| GCS on ED Arrival  13-15  9-12  3-8  Total  Median [IQR] | 69 (46.5%)  16 (7.9%)  82 (45.6%)*  167  9 [3,14] | 73 (19.8%)  83 (22.5%)  213 (57.7%)  369  7 [3, 12] | < 0.0001 |
| SAH  Yes  No  Total | 132 (79.0%)  35 (21.0%)  167 | 258 (74.1%)  90 (25.9%)  348 | 0.2243 |
| Scan Timing (days)  Median [IQR]  Total | 1 [1,2]  167 | 0 [0,1]  390 | < 0.0001 |
| Contusion Volume (cc)  Median [IQR]  Total | 2.3 [0.1, 18.7]  167 | 0.7 [0.04, 4.8]  390 | 0.0002 |
| * In TBIMS, participants who were chemically paralyzed or intubated did not receive a GCS score. They were assigned a score of GCS 3 for reporting. Chi-squared p-values are reported for categorical variables. | | | |

| Supplementary Table 4: 30cc Threshold Performance by Timepoint in TRACK-TBI | | | |
| --- | --- | --- | --- |
|  | TBIMS  (1 year)  N = 167 | TRACK-TBI  (6 month)  N = 340 | TRACK-TBI  (6 or 12 month)  N = 390 |
| AUC | 0.79 | 0.71 | 0.71 |
| Sensitivity | 0.67 | 0.24 | 0.16 |
| Specificity | 0.85 | 0.95 | 0.95 |
| Odds Ratio for contusions > 30 ccs [95% CI] | 11.5 [3.2, 41.3] | 4.1 [1.5, 11.2] | 5.9 [1.7, 20.1] |
| p value | 0.0002 | 0.006 | 0.005 |

| Supplementary Table 5: TRACK-TBI Epilepsy Cohort Demographics | | | |
| --- | --- | --- | --- |
|  | **No Epilepsy by 1-year**  **(N= 353)** | **Epilepsy by 1-year**  **(N = 37)** | p-value |
| Age (years)  Mean ± SD  Total | 37.6 ± 16.1  353 | 38.7 ± 15.3  37 | 0.5156 |
| Sex  Male  Female  Total | 267 (75.6%)  86 (24.4%)  353 | 30 (81.1%)  7 (18.9%)  37 | 0.547 |
| GCS on ED Arrival  13-15  9-12  3-8  Total | 68 (20.1%)  78 (23.1%)  192 (56.8%)*  338 | 5 (16.1%)  5 (16.1%)  21 (67.7%)  31 | 0.5558 |

| Supplementary Table 6: TBIMS and VHIS Cohort Demographics | | | |
| --- | --- | --- | --- |
|  | **TBI-Model Systems (Screening Cohort; N= 167)** | **VHIS**  **(External Validation Cohort; N=161)** | p-value |
| Age at Injury (years)  Mean ± SD  Total | 53.9 ± 22.4  166 | 21.2 ± 2.8  161 | < 0.0001 |
| Sex  Male  Female  Total | 113 (67.7%)  54 (32.3%)  167 | 161 (100%)  0 (0%)  161 | < 0.0001 |
| Race  White  Black  Other  Total | 133 (82.6%)  10 (6.2%)  18 (11.1%)  161 | 145 (90.0%)  13 (9.0%)  3 (6.0%)  161 | 0.003 |
| Ethnicity  Hispanic  Non-Hispanic  Total | 13 (7.8%)  154 (92.2%)  167 | 7 (4.3%)  154 (95.7%)  161 | 0.1935 |
| Education, years  Median [IQR]  Total | 14.5 [12, 16]  166 | 14 [12.8, 16]  159 | 0.054 |
| Loss of Consciousness  No  < 1 day  > 1 day  Total |  | 74 (46.8%)  68 (43.0%)  16 (10.1%)  158 |  |
| Contusion Volume (cc)  Median [IQR]  Total | 2.3 [0.1, 18.7]  167 | 31.0 [16.2, 57.1]  161 | < 0.0001 |

| Supplementary Table 7: VHIS Epilepsy Cohort Demographics | | | |
| --- | --- | --- | --- |
|  | **No Epilepsy by 1-year**  **(N= 88)** | **Epilepsy by 1-year**  **(N = 73)** | p-value |
| Age (years)  Mean ± SD  Total | 21.6 ± 3.3  88 | 20.7 ± 2.0  73 | 0.0987 |
| Sex  Male  Female  Total | 88 (100.0%)  0 (0.0%)  88 | 73 (100.0%)  0 (0.0%)  73 | 1 |
| Loss of Consciousness  No  < 1 day  > 1 day  Total | 43 (49.4%)  34 (39.1%)  10 (11.5%)*  87 | 31 (43.7%)  34 (47.9%)  6 (8.5%)  71 | 0.5119 |

**Supplementary Figures**

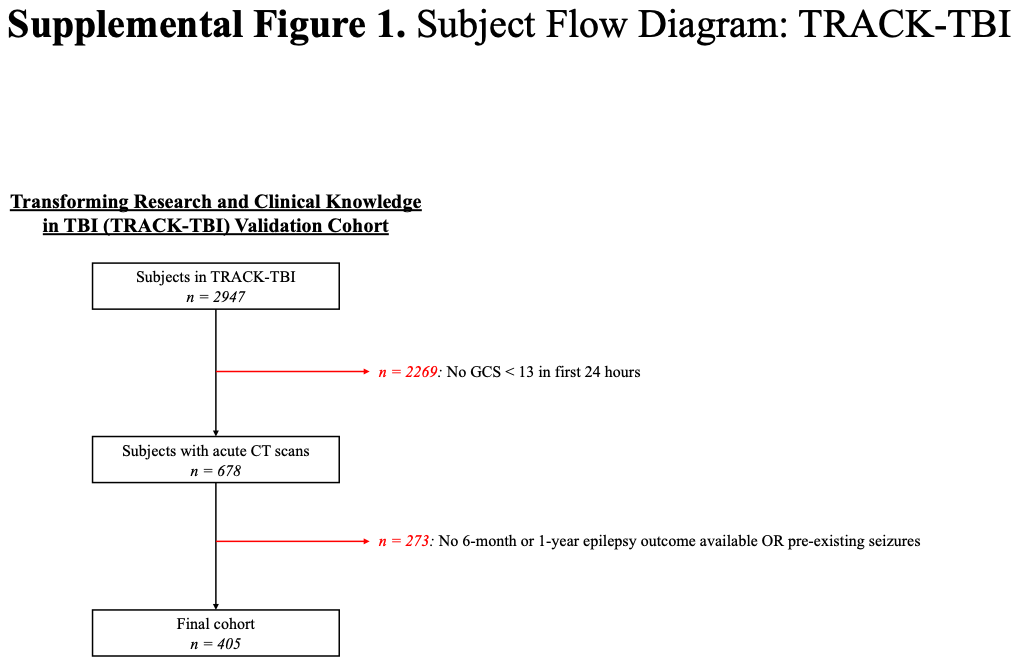

**Supplementary Figure 1** **CONSORT Diagram for TRACK-TBI.**

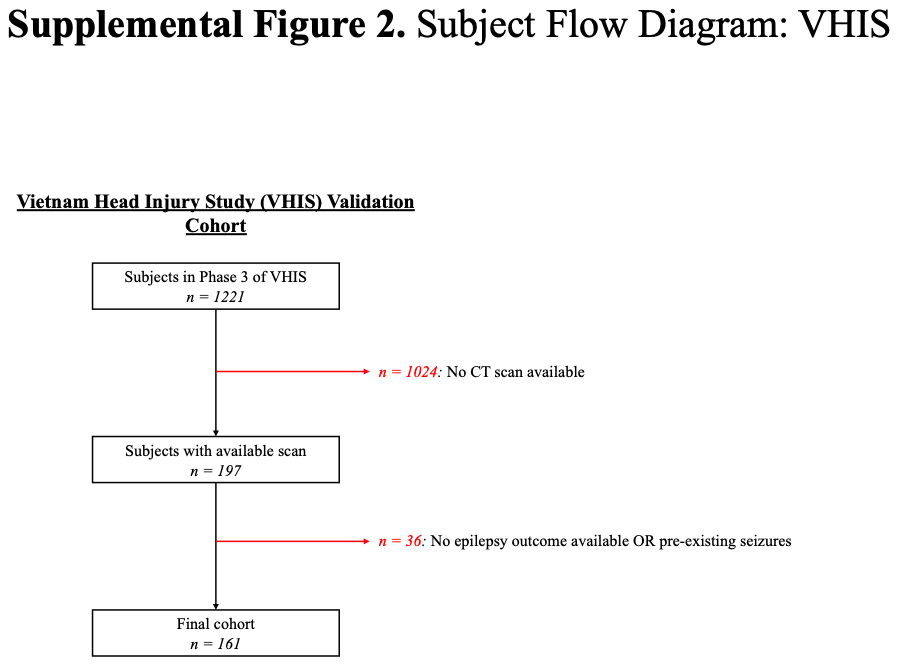

**Supplementary Figure 2** **CONSORT Diagram for VHIS.**

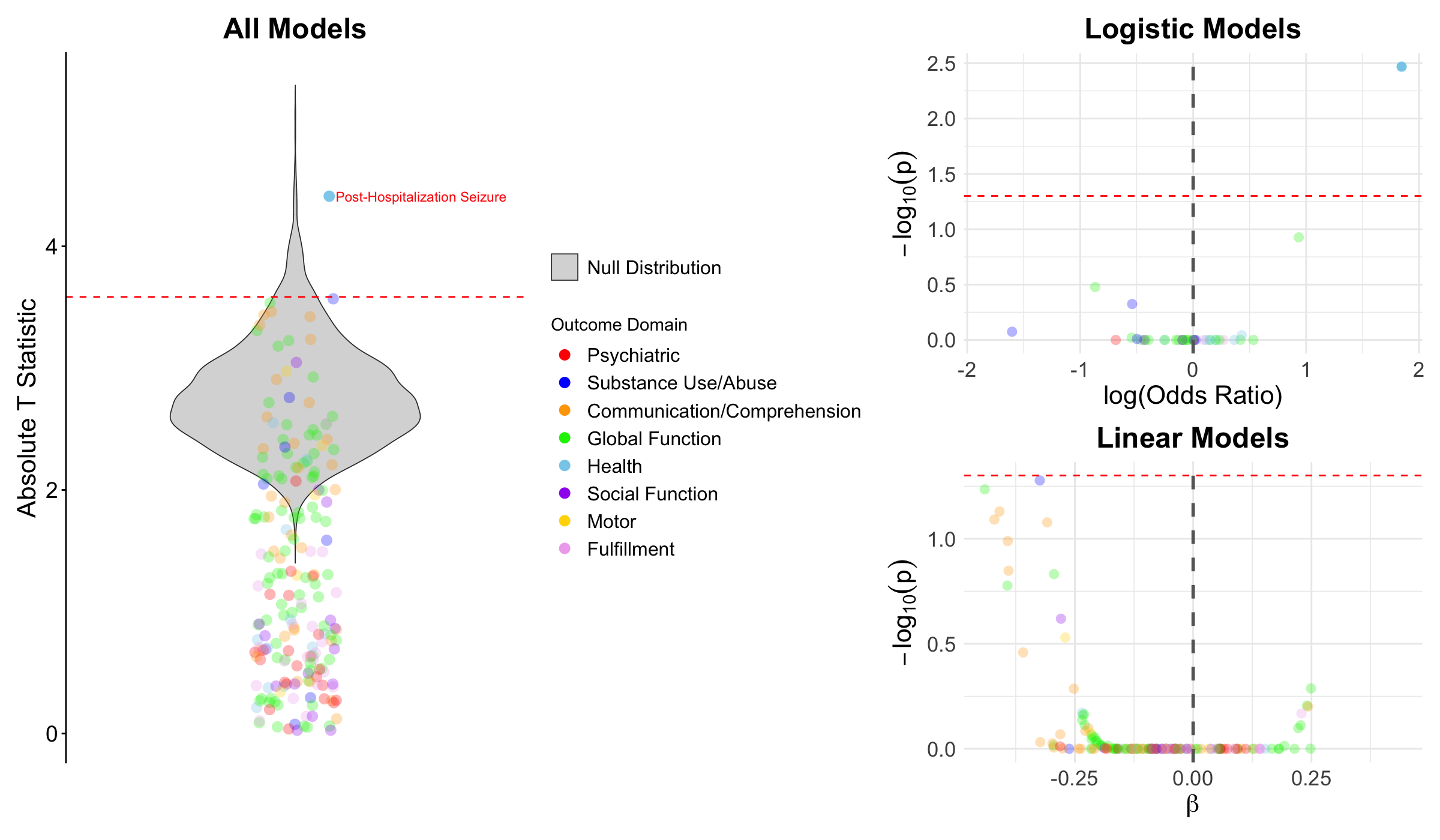

**Supplemental Figure 3 Regression Screen**

Each outcome in TBIMS was also assessed for association with contusion volume via either a logistic or linear regression. To provide a comparable statistical measure across the regression types, the absolute T-statistic was tested against a global null distribution.

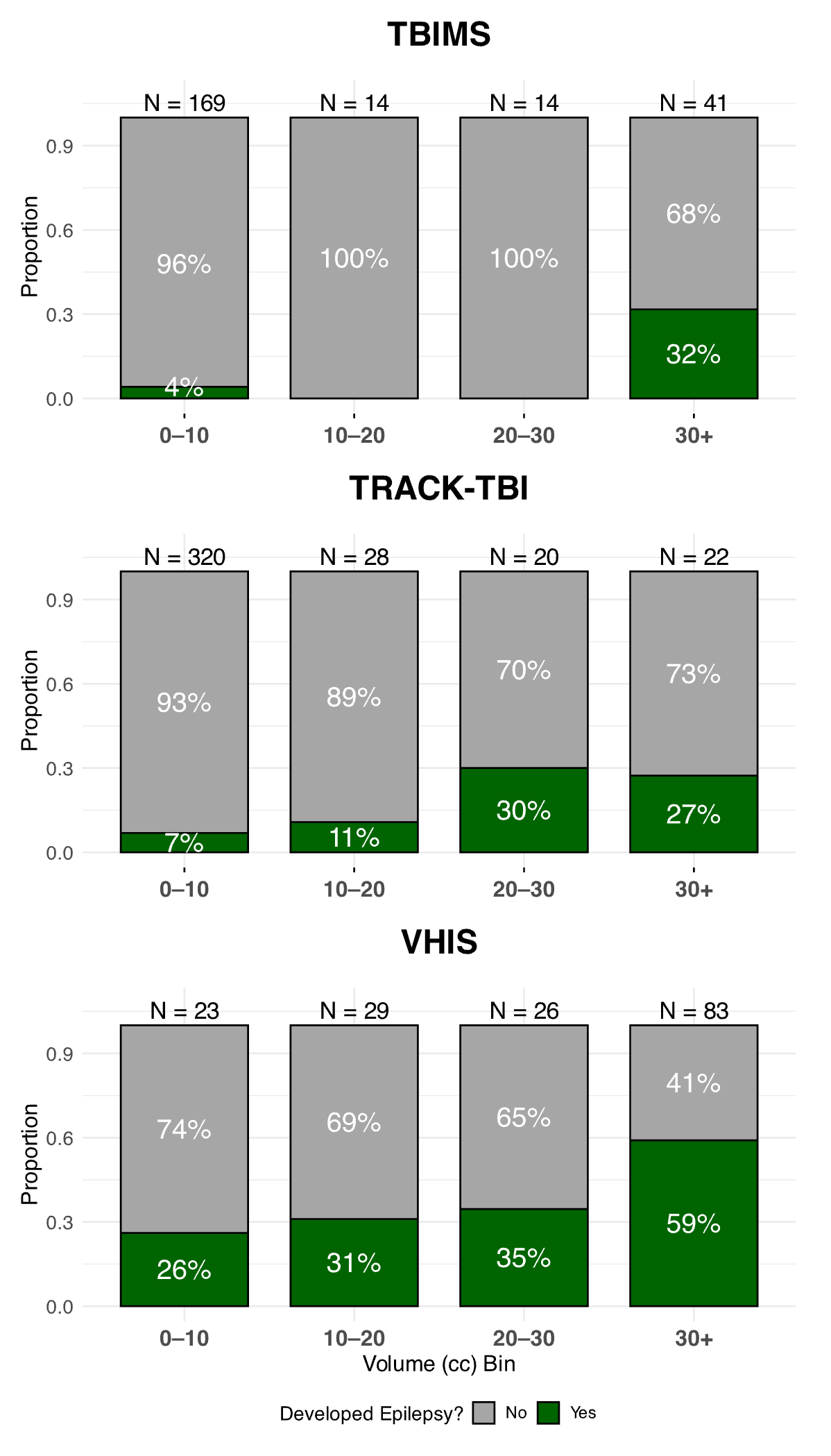

**Supplemental Figure 4 Epilepsy Proportion by Volume Bins**

In each cohort, the proportion of epilepsy was plotted in smaller 10 cc volume buckets to investigate whether epilepsy risk increased monotonically with contusion volume or the threshold indicated an inflection point.

**TBIMS Variable Appendix**

- ALCAnyDrinkF: During the past month have you had at least one drink of any alcoholic beverage such as beer, wine, wine coolers, or liquor?
  - 0 - No
  - 1 - Yes
- ALCDrinksF: A drink is 1 can or bottle of beer, 1 glass of wine, 1 can or bottle of wine cooler, 1 cocktail, or 1 shot of liquor. On the days when you drank, about how many drinks did you drink on average?
  - #
- ALCWeekF: During the past month, how many days per week did you drink any alcoholic beverages on the average?
  - #
- B3TCOMPF: Brief Test of Adult Cognition by Telephone (BTACT) - BTACT Total score standardized by age, sex and education
  - #
- B3TEFF: Brief Test of Adult Cognition by Telephone (BTACT) - BTACT executive functioning subscale
  - #
- B3TEMF: Brief Test of Adult Cognition by Telephone (BTACT) - BTACT episodic memory subscale
  - #
- BackCountDigitsF: Brief Test of Adult Cognition by Telephone (BTACT) - Backward counting number of digits produced:
  - #
- BackCountDigitsF_i_n: Brief Test of Adult Cognition by Telephone (BTACT) - Backward counting number of digits produced: standardized by age, sex, and education
  - #
- BackCountErrorsF: Brief Test of Adult Cognition by Telephone (BTACT) - Backward counting number of errors:
  - #
- BackCountLastNumF: Brief Test of Adult Cognition by Telephone (BTACT) - Backward counting last number reached:
  - #
- BackDigitCorrectF: Brief Test of Adult Cognition by Telephone (BTACT) - Backward digit span highest level reached:
  - #
- BackDigitCorrectF_i_n: Brief Test of Adult Cognition by Telephone (BTACT) - Backward digit span highest level reached: standardized by age, sex, and education
  - #
- BMICatF: BMI Category
  - 1 - Very severely underweight
  - 2 - Severely underweight
  - 3 - Underweight
  - 4 - Normal
  - 5 - Overweight
  - 6 - Obese Class I
  - 7 - Obese Class II
  - 8 - Obese Class III
- BMIF: BMI at Followup
  - #
- CombinedDRSF: Disability Rating Scale (DRS) - CombinedDRS is a variable that creates a single value based on DRSF, DRS_PI, and DRS_PI_ORIG values. This value uses DRS_PI_ORIG if it exists; if not, then it uses DRS_PI. If both are missing, then it uses DRSF.
  - #
- DAYSTo1stEmpF: Days From Injury to Employment
  - #
- DelayWordRecallCorrectF: Brief Test of Adult Cognition by Telephone (BTACT) – Delayed word recall correct
  - #
- DelayWordRecallCorrectF_i_n: Brief Test of Adult Cognition by Telephone (BTACT) – Delayed word recall total correct: standardized by age, sex, and education
  - #
- DelayWordRecallIntF: Brief Test of Adult Cognition by Telephone (BTACT) – Delay word recall number of intrusions
  - #
- DelayWordRecallMiddleF: Brief Test of Adult Cognition by Telephone (BTACT) – Delay word recall middle correct
  - #
- DelayWordRecallPrimacyF: Brief Test of Adult Cognition by Telephone (BTACT) – Delay word recall primacy correct
  - #
- DelayWordRecallRecencyF: Brief Test of Adult Cognition by Telephone (BTACT) – Delay word recall recency correct
  - #
- DelayWordRecallRepF: Brief Test of Adult Cognition by Telephone (BTACT) – Delay word recall number of repetitions
  - #
- DRINKCatF: Calculated drinking category
  - 0 - Abstaining
  - 1 - Light
  - 2 - Moderate
  - 3 - Heavy
- DRS_PI_ORIGF: DRS Interview at Follow-Up Matched with Original DRS is the post-acute interview (DRS Interview at Follow-Up) with the motor item and a communication item scored back to the original intent.
  - #
- DRS_PIEmpF: Disability Rating Scale Post-acute Interview (DRS-PI) - Employment subdomain of DRS as defined by the post-acute interview.
  - 0 - Not Restricted
  - 1 - Selected Jobs: Competitive
  - 2 - Sheltered Workshop: Non-competitive
  - 3 - Not Employable
- DRS_PIF: Disability Rating Scale Post-acute Interview (DRS-PI) - The DRS-PI provides a structured interview for administration of the Disability Rating Scale (DRS) over the telephone. Except for cases with very severe limitations (eg, minimally conscious), the scoring algorithm for the DRS-PI results in a score that is comparable to the original DRS. However, the Motor item of the DRS was not included in the DRS-PI because almost all cases interviewed in the development of the DRS-PI obtained a zero response on this item. In addition, the scoring of the Communication item was altered so that no score above 2 can be obtained.
  - #
- DRS_PIFeedF: Disability Rating Scale Post-acute Interview (DRS-PI) - Feeding subdomain of DRS as defined by the post-acute interview.
  - 0 - Complete
  - 1 - Partial
  - 2 - Minimal
  - 3 - None
- DRS_PIFuncF: Disability Rating Scale Post-acute Interview (DRS-PI) - Function subdomain of DRS as defined by the post-acute interview.
  - 0 - Completely Independent
  - 1 - Independent in Special Environment
  - 2 - Mildly Dependent: Limited assistance
  - 3 - Moderately Dependent: Moderate assistance
  - 4 - Markedly Dependent: Assist all major activities, all times
  - 5 - Totally Dependent: 24 hour nursing care
- DRS_PIGroomF: Disability Rating Scale Post-acute Interview (DRS-PI) - Grooming subdomain of DRS as defined by the post-acute interview.
  - 0 - Complete
  - 1 - Partial
  - 2 - Minimal
  - 3 - None
- DRS_PIToiletF: Disability Rating Scale Post-acute Interview (DRS-PI) - Toilet subdomain of DRS as defined by the post-acute interview.
  - 0 - Oriented
  - 1 - Confused
  - 2 - Inappropriate
  - 3 - Incomprehensible
  - 4 - None
- DRS_PIVerF: Disability Rating Scale Post-acute Interview (DRS-PI) - Verbal subdomain of DRS as defined by the post-acute interview.
  - 0 - Complete
  - 1 - Partial
  - 2 - Minimal
  - 3 - None
- drs2_1F: Disability Rating Scale (DRS) 2.1 - Is the subject able to communicate with you in a way that you and others clearly understand?
  - 1 - No
  - 2 - Inconsistently
  - 3 - Consistently
- drs2_3F: Disability Rating Scale (DRS) 2.3 - Are you [they] able to give your [their] correct name, location, year, month, day, and time of day promptly when asked?
  - 1 - No
  - 2 - Sometimes
  - 3 - Yes But Takes More Than A Few Seconds
  - 4 - Yes
- drs3_1F: Disability Rating Scale (DRS) 3.1 - Are you [they] able to obey commands? For example, move finger, look up, close eyes, stick out tongue.
  - 1 - No
  - 2 - Inconsistently
  - 3 - Yes
- drs4_1F: Disability Rating Scale (DRS) 4.1 - Can you feed yourself independently or manage tube feedings appropriately without help or reminders?
  - 1 - No
  - 2 - Yes
- drs4_2F: Disability Rating Scale (DRS) 4.2 - Do you understand what eating or feeding utensils or equipment are for and how they should be used?
  - 1 - Never
  - 2 - Some Of The Time
  - 3 - Most Of The Time
  - 4 - Always
- drs4_3F: Disability Rating Scale (DRS) 4.3 - Do you know when meal or feeding times are?
  - 1 - Never
  - 2 - Some Of The Time
  - 3 - Most Of The Time
  - 4 - Always
- drs5_1F: Disability Rating Scale (DRS) 5.1 - Can you use the toilet or manage your bowel and bladder routine independently and appropriately without help or reminders?
  - 1 - No
  - 2 - Yes
- drs5_2F: Disability Rating Scale (DRS) 5.2 - Do you understand how to manage your clothing or special equipment when toileting or in bowel and bladder management?
  - 1 - Never
  - 2 - Some Of The Time
  - 3 - Most Of The Time
  - 4 - Always
- drs5_3F: Disability Rating Scale (DRS) 5.3 - Do you know when to use the toilet or to conduct bowel and bladder management?
  - 1 - Never
  - 2 - Some Of The Time
  - 3 - Most Of The Time
  - 4 - Always
- drs6_1F: Disability Rating Scale (DRS) 6.1 - Can you dress and groom yourself independently and appropriately or direct someone else in these activities without help or reminders?
  - 1 - No
  - 2 - Yes
- drs6_2F: Disability Rating Scale (DRS) 6.2 - Do you know how to bathe and wash?
  - 1 - Never
  - 2 - Some Of The Time
  - 3 - Most Of The Time
  - 4 - Always
- drs6_3F: Disability Rating Scale (DRS) 6.3 - Do you understand how to get dressed?
  - 1 - Never
  - 2 - Some Of The Time
  - 3 - Most Of The Time
  - 4 - Always
- drs6_4F: Disability Rating Scale (DRS) 6.4 - Can you start and finish these grooming activities without prompting?
  - 1 - Never
  - 2 - Some Of The Time
  - 3 - Most Of The Time
  - 4 - Always
- drs7_1F: Disability Rating Scale (DRS) 7.1 - Do you function completely independently? That is, you do not require any physical assistance, supervision, equipment, devices, or reminders for cognitive, social, behavioral, emotional, and physical function?
  - 1 - No
  - 2 - Yes
- drs7_2F: Disability Rating Scale (DRS) 7.2 - Do you REQUIRE special aids or equipment such as a brace, walker, wheelchair, memory notebook, day planner, verbal reminders, prompts, cues, or alarm watch because of a disability?
  - 1 - No
  - 2 - Yes
- drs7_3F: Disability Rating Scale (DRS) 7.3 - Do you require PHYSICAL assistance from another person to meet daily needs?
  - 1 - Never
  - 2 - Some Of The Time
  - 3 - Most Of The Time
  - 4 - Always
- drs7_4F: Disability Rating Scale (DRS) 7.4 - Do you require assistance from another person in tasks that require THINKING ABILITIES?
  - 1 - Never
  - 2 - Some Of The Time
  - 3 - Most Of The Time
  - 4 - Always
- drs7_5F: Disability Rating Scale (DRS) 7.5 - Do you require assistance from another person to manage EMOTIONS AND BEHAVIOR?
  - 1 - Never
  - 2 - Some Of The Time
  - 3 - Most Of The Time
  - 4 - Always
- drs7_6aF: Disability Rating Scale (DRS) 7.6a - Do you take care of some of your needs but also need a helper who is always close by?
  - 1 - No
  - 2 - Yes
- drs7_6bF: Disability Rating Scale (DRS) 7.6b - Do you need help with all major activities and the assistance of another person all the time?
  - 1 - No
  - 2 - Yes
- drs7_6cF: Disability Rating Scale (DRS) 7.6c - Do you need 24-hour care and are not able to help with your own care at all?
  - 1 - No
  - 2 - Yes
- drs8_1F: Disability Rating Scale (DRS) 8.1 - Can you function with complete independence in work or social situations?
  - 1 - Never
  - 2 - Some Of The Time
  - 3 - Most Of The Time
  - 4 - Always
- drs8_2F: Disability Rating Scale (DRS) 8.2 - Can you understand, remember, and follow directions?
  - 1 - Never
  - 2 - Some Of The Time
  - 3 - Most Of The Time
  - 4 - Always
- drs8_3F: Disability Rating Scale (DRS) 8.3 - Can you keep track of time, schedules and appointments?
  - 1 - Never
  - 2 - Some Of The Time
  - 3 - Most Of The Time
  - 4 - Always
- drs8_4F: Disability Rating Scale (DRS) 8.4 - How certain are you that you can perform in a wide variety of jobs of your choosing or manage a home independently or participate in school full-time?
  - 1 - Certain Or Very Certain I Cannot
  - 2 - Uncertain
  - 3 - Certain Or Very Certain I Can
- drs8_5F: Disability Rating Scale (DRS) 8.5 - How certain are you that you can be successful at work, school or in home management with some reduction in the work load or with other accommodations due to disabilities?
  - 1 - Certain Or Very Certain I Cannot
  - 2 - Uncertain
  - 3 - Certain Or Very Certain I Can
- drs8_6F: Disability Rating Scale (DRS) 8.6 - How certain are you that you can be successful at work, school or in home management but with limited choices in jobs or school courses due to disabilities?
  - 1 - Certain Or Very Certain I Cannot
  - 2 - Uncertain
  - 3 - Certain Or Very Certain I Can
- drs8_7F: Disability Rating Scale (DRS) 8.7 - How certain are you that you can be able to work at home or in a special setting like a sheltered workshop in which the work is very routine and there is very frequent supervision and support?
  - 1 - Certain Or Very Certain I Cannot
  - 2 - Uncertain
  - 3 - Certain Or Very Certain I Can
- DrugsF: During the last 12 months did you use any illicit or non-prescription drugs?
  - 0 - No
  - 1 - Yes
- EarnF: Total annual salary based on current job(s)
  - 1 - $9,999 or less
  - 2 - $10,000 - $19,999
  - 3 - $20,000 - $29,999
  - 4 - $30,000 - $39,999
  - 5 - $40,000 - $49,999
  - 6 - $50,000 - $59,999
  - 7 - $60,000 - $69,999
  - 8 - $70,000 - $79,999
  - 9 - $80,000 - $89,999
  - 10 - $90,000 - $99,999
  - 11 - $100,000 or More
- FIMBathF: Functional Independence Measure (FIM): Bathing
  - 1 - Total Assist
  - 2 - Maximal Assist
  - 3 - Moderate Assist
  - 4 - Minimal Assist
  - 5 - Supervision
  - 6 - Modified Independence
  - 7 - Complete Independence
- FIMBedTransF: Functional Independence Measure (FIM): Bed, chair, wheelchair transfers
  - 1 - Total Assist
  - 2 - Maximal Assist
  - 3 - Moderate Assist
  - 4 - Minimal Assist
  - 5 - Supervision
  - 6 - Modified Independence
  - 7 - Complete Independence
- FIMBladAccF: Functional Independence Measure (FIM): Bladder management - frequency of accidents
  - 1 - Five or More Accidents in the Past 7 Days
  - 2 - Four Accidents in the Past 7 days
  - 3 - Three Accident in the Past 7 days
  - 4 - Two Accidents in the Past 7 days
  - 5 - One Accident in the Past 7 days
  - 6 - No Accidents: Uses device
  - 7 - No Accidents
- FIMBladAsstF: Functional Independence Measure (FIM): Bladder management - level of assistance
  - 1 - Total Assist
  - 2 - Maximal Assist
  - 3 - Moderate Assist
  - 4 - Minimal Assist
  - 5 - Supervision
  - 6 - Modified Independence
  - 7 - Complete Independence
- FIMBladMgtF: Functional Independence Measure (FIM): Bladder management
  - 1 - Total Assist
  - 2 - Maximal Assist
  - 3 - Moderate Assist
  - 4 - Minimal Assist
  - 5 - Supervision
  - 6 - Modified Independence
  - 7 - Complete Independence
- FIMBwlAccF: Functional Independence Measure (FIM): Bowel management - frequency of accidents
  - 1 - Five or More Accidents in the Past 7 Days
  - 2 - Four Accidents in the Past 7 days
  - 3 - Three Accident in the Past 7 days
  - 4 - Two Accidents in the Past 7 days
  - 5 - One Accident in the Past 7 days
  - 6 - No Accidents: Uses device
  - 7 - No Accidents
- FIMBwlAsstF: Functional Independence Measure (FIM): Bowel management - level of assistance
  - 1 - Total Assist
  - 2 - Maximal Assist
  - 3 - Moderate Assist
  - 4 - Minimal Assist
  - 5 - Supervision
  - 6 - Modified Independence
  - 7 - Complete Independence
- FIMBwlMgtF: Functional Independence Measure (FIM): Bowel management
  - 1 - Total Assist
  - 2 - Maximal Assist
  - 3 - Moderate Assist
  - 4 - Minimal Assist
  - 5 - Supervision
  - 6 - Modified Independence
  - 7 - Complete Independence
- FIMCOGF: Functional Independence Measure (FIM): FIM Cognitive Follow-up
  - #
- FIMCompF: Functional Independence Measure (FIM): Comprehension
  - 1 - Total Assist
  - 2 - Maximal Assist
  - 3 - Moderate Assist
  - 4 - Minimal Assist
  - 5 - Supervision
  - 6 - Modified Independence
  - 7 - Complete Independence
- FIMDrsdwnF: Functional Independence Measure (FIM): Dressing lower body
  - 1 - Total Assist
  - 2 - Maximal Assist
  - 3 - Moderate Assist
  - 4 - Minimal Assist
  - 5 - Supervision
  - 6 - Modified Independence
  - 7 - Complete Independence
- FIMDrupF: Functional Independence Measure (FIM): Dressing upper body
  - 1 - Total Assist
  - 2 - Maximal Assist
  - 3 - Moderate Assist
  - 4 - Minimal Assist
  - 5 - Supervision
  - 6 - Modified Independence
  - 7 - Complete Independence
- FIMExpressF: Functional Independence Measure (FIM): Expression
  - 1 - Total Assist
  - 2 - Maximal Assist
  - 3 - Moderate Assist
  - 4 - Minimal Assist
  - 5 - Supervision
  - 6 - Modified Independence
  - 7 - Complete Independence
- FIMFeedF: Functional Independence Measure (FIM): Eating
  - 1 - Total Assist
  - 2 - Maximal Assist
  - 3 - Moderate Assist
  - 4 - Minimal Assist
  - 5 - Supervision
  - 6 - Modified Independence
  - 7 - Complete Independence
- FIMGroomF: Functional Independence Measure (FIM): Grooming
  - 1 - Total Assist
  - 2 - Maximal Assist
  - 3 - Moderate Assist
  - 4 - Minimal Assist
  - 5 - Supervision
  - 6 - Modified Independence
  - 7 - Complete Independence
- FIMLocoF: Functional Independence Measure (FIM): Locomotion: walk/wheelchair
  - 1 - Total Assist
  - 2 - Maximal Assist
  - 3 - Moderate Assist
  - 4 - Minimal Assist
  - 5 - Supervision
  - 6 - Modified Independence
  - 7 - Complete Independence
- FIMMemF: Functional Independence Measure (FIM): Memory
  - 1 - Total Assist
  - 2 - Maximal Assist
  - 3 - Moderate Assist
  - 4 - Minimal Assist
  - 5 - Supervision
  - 6 - Modified Independence
  - 7 - Complete Independence
- FIMMOTF: Functional Independence Measure (FIM): FIM Motor Followup
  - #
- FIMProbSlvF: Functional Independence Measure (FIM): Problem solving
  - 1 - Total Assist
  - 2 - Maximal Assist
  - 3 - Moderate Assist
  - 4 - Minimal Assist
  - 5 - Supervision
  - 6 - Modified Independence
  - 7 - Complete Independence
- FIMSocialF: Functional Independence Measure (FIM): Social interaction
  - 1 - Total Assist
  - 2 - Maximal Assist
  - 3 - Moderate Assist
  - 4 - Minimal Assist
  - 5 - Supervision
  - 6 - Modified Independence
  - 7 - Complete Independence
- FIMStairsF: Functional Independence Measure (FIM): Stairs
  - 1 - Total Assist
  - 2 - Maximal Assist
  - 3 - Moderate Assist
  - 4 - Minimal Assist
  - 5 - Supervision
  - 6 - Modified Independence
  - 7 - Complete Independence
- FIMToiletF: Functional Independence Measure (FIM): Toileting
  - 1 - Total Assist
  - 2 - Maximal Assist
  - 3 - Moderate Assist
  - 4 - Minimal Assist
  - 5 - Supervision
  - 6 - Modified Independence
  - 7 - Complete Independence
- FIMToilTransF: Functional Independence Measure (FIM): Toilet transfers
  - 1 - Total Assist
  - 2 - Maximal Assist
  - 3 - Moderate Assist
  - 4 - Minimal Assist
  - 5 - Supervision
  - 6 - Modified Independence
  - 7 - Complete Independence
- FIMTOTF: Functional Independence Measure (FIM): FIM Total (New) Follow-up
  - #
- FIMTubTransF: Functional Independence Measure (FIM): Tub or shower transfers
  - 1 - Total Assist
  - 2 - Maximal Assist
  - 3 - Moderate Assist
  - 4 - Minimal Assist
  - 5 - Supervision
  - 6 - Modified Independence
  - 7 - Complete Independence
- FluencyCorrect1_15F: Brief Test of Adult Cognition by Telephone (BTACT) - Category fluency total correct 1 - 15 sec
  - #
- FluencyCorrect15_30F: Brief Test of Adult Cognition by Telephone (BTACT) - Category fluency total correct 15 - 30 sec
  - #
- FluencyCorrect30_45F: Brief Test of Adult Cognition by Telephone (BTACT) - Category fluency total correct 30 - 45 sec
  - #
- FluencyCorrect45_60F: Brief Test of Adult Cognition by Telephone (BTACT) - Category fluency total correct 45 - 60 sec
  - #
- FluencyCorrectF: Brief Test of Adult Cognition by Telephone (BTACT) - Category fluency total correct:
  - #
- FluencyCorrectF_i_n: Brief Test of Adult Cognition by Telephone (BTACT) - Category fluency total correct: standardized by age, sex, and education
  - #
- FluencyRepF: Brief Test of Adult Cognition by Telephone (BTACT) - Category fluency number of repetitions
  - #
- GAD7TOTF: Generalized Anxiety Disorder Scale (GAD) - Generalized Anxiety Disorder Total Score
  - #
- GADAfraidF: Generalized Anxiety Disorder Scale (GAD) g. - Over the LAST 2 WEEKS, how often have you been bothered by: Feeling afraid as if something awful might happen
  - 0 - Not at All
  - 1 - Several Days
  - 2 - More Than Half of the Days
  - 3 - Nearly Every day
- GADAnnoyF: Generalized Anxiety Disorder Scale (GAD) f. - Over the LAST 2 WEEKS, how often have you been bothered by: Becoming easily annoyed or irritable
  - 0 - Not at All
  - 1 - Several Days
  - 2 - More Than Half of the Days
  - 3 - Nearly Every day
- GADCntrlWryF: Generalized Anxiety Disorder Scale (GAD) b. - Over the LAST 2 WEEKS, how often have you been bothered by: Not being able to stop or control worrying
  - 0 - Not at All
  - 1 - Several Days
  - 2 - More Than Half of the Days
  - 3 - Nearly Every day
- GADDifficultF: Generalized Anxiety Disorder Scale (GAD) h. - Over the LAST 2 WEEKS, how difficult have these problems made it for you to do your work, take care of things at home, or get along with other people?
  - 0 - Not Difficult at All
  - 1 - Somewhat Difficult
  - 2 - Very Difficult
  - 3 - Extremely Difficult
- GADNervousF: Generalized Anxiety Disorder Scale (GAD) a. - Over the LAST 2 WEEKS, how often have you been bothered by: Feeling nervous, anxious or on edge
  - 0 - Not at All
  - 1 - Several Days
  - 2 - More Than Half of the Days
  - 3 - Nearly Every day
- GADRelaxF: Generalized Anxiety Disorder Scale (GAD) d. - Over the LAST 2 WEEKS, how often have you been bothered by: Trouble relaxing
  - 0 - Not at All
  - 1 - Several Days
  - 2 - More Than Half of the Days
  - 3 - Nearly Every day
- GADRestlessF: Generalized Anxiety Disorder Scale (GAD) e. - Over the LAST 2 WEEKS, how often have you been bothered by: Being so restless that it is hard to sit still
  - 0 - Not at All
  - 1 - Several Days
  - 2 - More Than Half of the Days
  - 3 - Nearly Every day
- GADWorryF: Generalized Anxiety Disorder Scale (GAD) c. - Over the LAST 2 WEEKS, how often have you been bothered by: Worrying too much about different things
  - 0 - Not at All
  - 1 - Several Days
  - 2 - More Than Half of the Days
  - 3 - Nearly Every day
- GenHlthF: In general would you say your health is ...
  - 1 - Excellent
  - 2 - Very Good
  - 3 - Good
  - 4 - Fair
  - 5 - Poor
- GOSAssistAllF: Glasgow Outcome Scale (GOS) 2a. - Is the assistance of another person at home essential every day for some activities of daily living?
  - 0 - No
  - 1 - Yes
- GOSDisruptF: Glasgow Outcome Scale (GOS) 7a. - Have there been psychological problems which have resulted in ongoing family disruption or disruption to friendships?
  - 0 - No
  - 1 - Yes
- GOSEF - Glasgow Outcome Scale - Extended (GOS-E) - GOS-E Incl. Expired
  - 1 - Dead
  - 2 - Vegetative State (VS)
  - 3 - Lower Severe Disability (LSD)
  - 4 - Upper Severe Disability (USD)
  - 5 - Lower Moderate Disability (LMD)
  - 6 - Upper Moderate Disability (UMD)
  - 7 - Lower Good Recovery (LGR)
  - 8 - Upper Good Recovery (UGR)
- GOSEF_bin: Glasgow Outcome Scale - Extended (GOS-E) - GOS-E score binarized at 4
  - 0 - GOSEF score of 5, 6, 7, 8
  - 1 - GOSEF score of 1, 2, 3, 4
- GOSFrqHlpF: Glasgow Outcome Scale (GOS) 2b. - Do you need frequent help or someone to be around at home most of the time?
  - 0 - No
  - 1 - Yes
- GOSPrbCurrentF: Glasgow Outcome Scale (GOS) 8a - Are there any other current problems relating to the injury which affect daily life?
  - 0 - No
  - 1 - Yes
- GOSShopF: Glasgow Outcome Scale (GOS) 3a. - Are you able to shop without assistance?
  - 0 - No
  - 1 - Yes
- GOSSocF: Glasgow Outcome Scale (GOS) 6a. - Are you able to resume regular social and leisure activities outside home?
  - 0 - No
  - 1 - Yes
- GOSTotalF: Glasgow Outcome Scale (GOS) 9. - GOS-E score:
  - 1 - Dead
  - 2 - Vegetative State (VS)
  - 3 - Lower Severe Disability (LSD)
  - 4 - Upper Severe Disability (USD)
  - 5 - Lower Moderate Disability (LMD)
  - 6 - Upper Moderate Disability (UMD)
  - 7 - Lower Good Recovery (LGR)
  - 8 - Upper Good Recovery (UGR)
- GOSTravelF: Glasgow Outcome Scale (GOS) 4a. - Are you able to travel locally without assistance?
  - 0 - No
  - 1 - Yes
- GOSWorkF: Glasgow Outcome Scale (GOS) 5a. - Are you currently able to work to your previous capacity?
  - 0 - No
  - 1 - Yes
- HighBloodCholesterol_New: In patients with no history of high blood cholesterol prior to injury, did they report having a new onset of high blood cholesterol at the same time or after their injury?
  - 0 - No
  - 1 - Yes
- Hypertension_New: In patients with no history of high blood cholesterol prior to injury, did they report having a new onset of high blood cholesterol at the same time or after their injury?
  - 0 - No
  - 1 - Yes
- Malec_EatOutF: Participation Assessment with Recombined Tools-Objective (PART-O) - Rasch modified scoring for PART-O: Part Malec eat out Score
  - #
- Malec_FriendF: Participation Assessment with Recombined Tools-Objective (PART-O) - Rasch modified scoring for PART-O: Part Malec friend Score
  - #
- Malec_MovieF: Participation Assessment with Recombined Tools-Objective (PART-O) - Rasch modified scoring for PART-O: Part Malec movie Score
  - #
- Malec_OutHseF: Participation Assessment with Recombined Tools-Objective (PART-O) - Rasch modified scoring for PART-O: Part Malec out-of-house Score
  - #
- Malec_PlaySportF: Participation Assessment with Recombined Tools-Objective (PART-O) - Rasch modified scoring for PART-O: Part Malec engaging in sports Score
  - #
- Malec_ProdF: Participation Assessment with Recombined Tools-Objective (PART-O) - Rasch modified scoring for PART-O: Part Malec productivity Score
  - #
- Malec_RelationF: Participation Assessment with Recombined Tools-Objective (PART-O) - Rasch modified scoring for PART-O: Part Malec relationship Score
  - #
- Malec_ReligionF: Participation Assessment with Recombined Tools-Objective (PART-O) - Rasch modified scoring for PART-O: Part Malec religion Score
  - #
- Malec_ShopF: Participation Assessment with Recombined Tools-Objective (PART-O) - Rasch modified scoring for PART-O: Part Malec shopping Score
  - #
- Malec_SocialF: Participation Assessment with Recombined Tools-Objective (PART-O) - Rasch modified scoring for PART-O: Part Malec social Score
  - #
- Malec_SumF: Participation Assessment with Recombined Tools-Objective (PART-O) - Rasch modified scoring for PART-O: Part Malec summary Score
  - #
- Malec_WatchSportF: Participation Assessment with Recombined Tools-Objective (PART-O) - Rasch modified scoring for PART-O: Part Malec watching sports Score
  - #
- MJUseF: Did you use marijuana?
  - 0 - No
  - 1 - Yes
- PanicAttacks_New: In patients with no history of panic attacks prior to injury, did they report having a new onset of panic attacks at the same time or after their injury?
  - 0 - No
  - 1 - Yes
- PART_BalancedF: Participation Assessment with Recombined Tools-Objective (PART-O) - Weighted PART Score
  - #
- PART_Domain_OutF: Participation Assessment with Recombined Tools-Objective (PART-O) - Weighted Out and About PART Score
  - #
- PART_Domain_ProdF: Participation Assessment with Recombined Tools-Objective (PART-O) - Weighted Productivity PART Score
  - #
- PART_Domain_SocF: Participation Assessment with Recombined Tools-Objective (PART-O) - Weighted Social PART Score
  - #
- PART_RaschF: Participation Assessment with Recombined Tools-Objective (PART-O) - Rasch PART Score
  - #
- PART_SDF: Participation Assessment with Recombined Tools-Objective (PART-O) - Weighted PART Standardized Deviation Score
  - #
- PARTOutAboutF: Participation Assessment with Recombined Tools-Objective (PART-O) - Part OutAbout Subscale
  - #
- PARTProductivityF: Participation Assessment with Recombined Tools-Objective (PART-O) - Part Productivity Subscale
  - #
- PARTSocialF: Participation Assessment with Recombined Tools-Objective (PART-O) - Part Social Subscale
  - #
- PARTSummaryF: Participation Assessment with Recombined Tools-Objective (PART-O) - Part Summary Statistic
  - #
- PastYearSeizF: How many seizures have you had in the past year (or since your discharge)?
  - 1 - up to three seizures
  - 2 - 4-12 seizures
  - 3 - at least one seizure monthly
  - 4 - at least one seizure weekly
  - 5 - at least one seizure daily
  - 88 - Not applicable: No seizures
- PHQ9TOTF: Patient Health Questionnaire (PHQ) - Sum of nine depression items total score
  - #
- PHQBadF: Patient Health Questionnaire-2 (PHQ-2) - Over the LAST 2 WEEKS, how often have you been bothered by: Feeling bad about yourself or that you are a failure or have let yourself or your family down
  - 0 - Not at All
  - 1 - Several Days
  - 2 - More Than Half of the Days
  - 3 - Nearly Every day
- PHQConcentrateF: Patient Health Questionnaire-2 (PHQ-2) - Over the LAST 2 WEEKS, how often have you been bothered by: Trouble concentrating on things, such as reading the newspaper or watching television
  - 0 - Not at All
  - 1 - Several Days
  - 2 - More Than Half of the Days
  - 3 - Nearly Every day
- PHQDeadF: Patient Health Questionnaire-2 (PHQ-2) - Over the LAST 2 WEEKS, how often have you been bothered by: Thoughts that you would be better off dead or hurting yourself in some way
  - 0 - Not at All
  - 1 - Several Days
  - 2 - More Than Half of the Days
  - 3 - Nearly Every day
- PHQDifficultF: Patient Health Questionnaire-2 (PHQ-2) - Over the LAST 2 WEEKS, how often have you been bothered by: How difficult have these problems made it for you to do your work, take care of things at home, or get along with other people?
  - 0 - Not Difficult at All
  - 1 - Somewhat Difficult
  - 2 - Very Difficult
  - 3 - Extremely Difficult
- PHQDownF: Patient Health Questionnaire-2 (PHQ-2) - Over the LAST 2 WEEKS, how often have you been bothered by: Feeling down, depressed, or hopeless
  - 0 - Not at All
  - 1 - Several Days
  - 2 - More Than Half of the Days
  - 3 - Nearly Every day
- PHQEAtF: Patient Health Questionnaire-2 (PHQ-2) - Over the LAST 2 WEEKS, how often have you been bothered by: Poor appetite or overeating
  - 0 - Not at All
  - 1 - Several Days
  - 2 - More Than Half of the Days
  - 3 - Nearly Every day
- PHQPleasureF: Patient Health Questionnaire-2 (PHQ-2) – Over the LAST 2 WEEKS, how often have you been bothered by: Little interest or pleasure in doing things
  - 0 - Not at All
  - 1 - Several Days
  - 2 - More Than Half of the Days
  - 3 - Nearly Every day
- PHQSleepF: Patient Health Questionnaire-2 (PHQ-2) - Over the LAST 2 WEEKS, how often have you been bothered by: Trouble falling or staying asleep, or sleeping too much
  - 0 - Not at All
  - 1 - Several Days
  - 2 - More Than Half of the Days
  - 3 - Nearly Every day
- PHQSlowF: Patient Health Questionnaire-2 (PHQ-2) - Over the LAST 2 WEEKS, how often have you been bothered by: Moving or speaking so slowly that other people could have noticed. Or the opposite - being so fidgety or restless that you have been moving around a lot more than usual
  - 0 - Not at All
  - 1 - Several Days
  - 2 - More Than Half of the Days
  - 3 - Nearly Every day
- PHQTiredF: Patient Health Questionnaire-2 (PHQ-2) - Over the LAST 2 WEEKS, how often have you been bothered by: Feeling tired or having little energy
  - 0 - Not at All
  - 1 - Several Days
  - 2 - More Than Half of the Days
  - 3 - Nearly Every day
- PhysHlthF: Compared to (after your discharge from the rehab center) or (one year ago), how would you rate your physical health in general now?
  - 1 - Excellent
  - 2 - Very Good
  - 3 - Good
  - 4 - Fair
  - 5 - Poor
- PROBLEMUseF: Substance Problem Use defined as either taken illicit drugs, binge drinking (5 or more drinks on occasion) in the past month, or their drinking category is Heavy
  - 0 - No
  - 1 - Yes
- PRTEatOutF: Participation Assessment with Recombined Tools-Objective (PART-O) - In a typical month, how many times do you eat in a restaurant?
  - 0 - None
  - 1 - 1 - 4 Times
  - 2 - 5 - 9 Times
  - 3 - 10 - 19 Times
  - 4 - 20 - 34 Times
  - 5 - 35 or More Times
- PRTEmotSupF: Participation Assessment with Recombined Tools-Objective (PART-O) - In a typical week, how many times do you give emotional support to other people, that is, listen to their problems or help them with their troubles?
  - 0 - None
  - 1 - 1 - 4 Times
  - 2 - 5 - 9 Times
  - 3 - 10 - 19 Times
  - 4 - 20 - 34 Times
  - 5 - 35 or More Times
- PRTFriendF: Participation Assessment with Recombined Tools-Objective (PART-O) – Not including your spouse or significant other, do you have a close friend in whom you confide?
  - 0 - No
  - 1 - Yes
- PRTHomeF: Participation Assessment with Recombined Tools-Objective (PART-O) - In a typical week, how many hours do you spend in active homemaking, including cleaning, cooking and raising children?
  - 0 - None
  - 1 - 1 - 4 Hours
  - 2 - 5 - 9 Hours
  - 3 - 10 - 19 Hours
  - 4 - 20 - 34 Hours
  - 5 - 35 or More Hours
- PRTInternetF: Participation Assessment with Recombined Tools-Objective (PART-O) - In a typical week, how many times do you use the Internet for communication with others? For example, text, email, virtual meetings, social media?
  - 0 - None
  - 1 - 1 - 4 Times
  - 2 - 5 - 9 Times
  - 3 - 10 - 19 Times
  - 4 - 20 - 34 Times
  - 5 - 35 or More Times
- PRTMovieF: Participation Assessment with Recombined Tools-Objective (PART-O) - In a typical month, how many times do you go to the movies?
  - 0 - None; 1 - One Time
  - 2 - Two Times
  - 3 - Three Times
  - 4 - Four Times
  - 5 - Five or More Times
- PRTOutHseF: Participation Assessment with Recombined Tools-Objective (PART-O) - In a typical week, how many days do you get out of your house and go somewhere? It could be anywhere. It doesn't have to be any place "special".
  - 0 - None
  - 1 - 1 - 2 Days
  - 2 - 3 - 4 Days
  - 3 - 5 - 6 Days
  - 4 - 7 Days
- PRTPlaySportF: Participation Assessment with Recombined Tools-Objective (PART-O) - In a typical month, how many times do you engage in sports or exercise outside your home? Include activities like running, bowling, going to the gym, swimming, walking for exercise and the like.
  - 0 - None
  - 1 - 1 - 4 Times
  - 2 - 5 - 9 Times
  - 3 - 10 - 19 Times
  - 4 - 20 - 34 Times
  - 5 - 35 or More Times
- PRTRelationF: Participation Assessment with Recombined Tools-Objective (PART-O) – Are you currently involved in an ongoing intimate, that is, romantic or sexual, relationship??
  - 0 - No
  - 1 - Yes
- PRTReligionF: Participation Assessment with Recombined Tools-Objective (PART-O) - In a typical month, how many times do you do attend religious or spiritual services? Include places like churches, temples and mosques.
  - 0 - None; 1 - One Time
  - 2 - Two Times
  - 3 - Three Times
  - 4 - Four Times
  - 5 - Five or More Times
- PRTSchoolF: Participation Assessment with Recombined Tools-Objective (PART-O) - In a typical week, how many hours do you spend in school working toward a degree or in an accredited technical training program, including hours in class and studying?
  - 0 - None
  - 1 - 1 - 4 Hours
  - 2 - 5 - 9 Hours
  - 3 - 10 - 19 Hours
  - 4 - 20 - 34 Hours
  - 5 - 35 or More Hours
- PRTShopF: Participation Assessment with Recombined Tools-Objective (PART-O) - In a typical month, how many times do you go shopping? Include grocery shopping, as well as shopping for household necessities, or just for fun.
  - 0 - None
  - 1 - 1 - 4 Times
  - 2 - 5 - 9 Times
  - 3 - 10 - 19 Times
  - 4 - 20 - 34 Times
  - 5 - 35 or More Times
- PRTSocFamF: Participation Assessment with Recombined Tools-Objective (PART-O) - In a typical week, how many times do you socialize with family and relatives, in person or by phone?
  - 0 - None
  - 1 - 1 - 4 Times
  - 2 - 5 - 9 Times
  - 3 - 10 - 19 Times
  - 4 - 20 - 34 Times
  - 5 - 35 or More Times
- PRTSocFrndF: Participation Assessment with Recombined Tools-Objective (PART-O) - In a typical week, how many times do you socialize with friends, in person or by phone?
  - 0 - None
  - 1 - 1 - 4 Times
  - 2 - 5 - 9 Times
  - 3 - 10 - 19 Times
  - 4 - 20 - 34 Times
  - 5 - 35 or More Times
- PRTSpouseF: Participation Assessment with Recombined Tools-Objective (PART-O) – Do you live with your spouse or significant other?
  - 0 - No
  - 1 - Yes
- PRTVolF: Participation Assessment with Recombined Tools-Objective (PART-O) - In a typical month, how many times do you do volunteer work?
  - 0 - None
  - 1 - One Time
  - 2 - Two Times
  - 3 - Three Times
  - 4 - Four Times
  - 5 - Five or More Times
- PRTWorkF: Participation Assessment with Recombined Tools-Objective (PART-O) - In a typical week, how many hours do you spend working for money, whether in a job or self-employed?
  - 0 - None
  - 1 - 1 - 4 Hours
  - 2 - 5 - 9 Hours
  - 3 - 10 - 19 Hours
  - 4 - 20 - 34 Hours
  - 5 - 35 or More Hours
- PRTWtchSportF: Participation Assessment with Recombined Tools-Objective (PART-O) - In a typical month, how many times do you attend sports events in person, as a spectator?
  - 0 - None; 1 - One Time
  - 2 - Two Times
  - 3 - Three Times
  - 4 - Four Times
  - 5 - Five or More Times
- Reason01F: Brief Test of Adult Cognition by Telephone (BTACT) - Reasoning Trial 1
  - 0 - Incorrect
  - 1 - Correct
- Reason02F: Brief Test of Adult Cognition by Telephone (BTACT) - Reasoning Trial 2
  - 0 - Incorrect
  - 1 - Correct
- Reason03F: Brief Test of Adult Cognition by Telephone (BTACT) - Reasoning Trial 3
  - 0 - Incorrect
  - 1 - Correct
- Reason04F: Brief Test of Adult Cognition by Telephone (BTACT) - Reasoning Trial 4
  - 0 - Incorrect
  - 1 - Correct
- Reason05F: Brief Test of Adult Cognition by Telephone (BTACT) - Reasoning Trial 1
  - 0 - Incorrect
  - 1 - Correct
- ReasonCorrectF: Brief Test of Adult Cognition by Telephone (BTACT) - Reasoning total correct
  - #
- ReasonCorrectF_i_n: Brief Test of Adult Cognition by Telephone (BTACT) -Reasoning total correct: standardized by age, sex, and education
  - #
- REHOSPF: Rehospitalized in Past Year
  - 0 - No
  - 1 - Yes
- SRScal2F: Supervision Rating Scale 1
  - 1 - Independent
  - 2 - Overnight or Part Time Supervision
  - 3 - Full Time Indirect or Direct Supervision
- SRScalF: Supervision Rating Scale 2
  - 2 - Independent
  - 3 - The Person with Brain Injury is Only Supervised Overnight ( One or more supervising persons are always present overnight but they are all sometimes absent for the rest of the day. )
  - 4 - The Person with Brain Injury is Supervised Overnight and Selected Day Times ( But is allowed on independent outings. )
  - 5 - The Person with Brain Injury is Supervised Overnight and Part Time During Day Times ( But unsupervised during working hours )
  - 6 - The Person with Brain Injury is Supervised Overnight and During Most Waking Hours ( Supervising persons are all sometimes absent for periods longer than one hour, but less than the time needed to hold a full-time job away from home. )
  - 7 - The Person with Brain Injury is Supervised Overnight and During Almost All Waking Hours ( Persons are all sometimes absent for periods shorter than one hour. )
  - 8 - The Person with Brain Injury is Under Full Time Indirect Supervision ( At least one supervising person is always present, but the supervising person does not check on the person with brain injury more than once every 30 minutes )
  - 9 - Same as #8 but Requires Overnight Safety Precautions ( A deadbolt on outside door )
  - 10 - The Person with Brain Injury is Under Full Time Direct Supervision ( At least one supervising person is always present and the supervising person checks on the person with brain injury more than once every thirty minutes )
  - 11 - The Person Lives in Home where the Exits are Physically Controlled ( A locked ward )
  - 12 - Same as #11 but a Supervising Person is Designated to Provide Full Time Line-of-Sight Supervision ( An escape watch or suicide watch )
  - 13 - The Person with Brain Injury is in Physical Restraints
- Stroke_New: In patients with no history of stroke prior to injury, did they report having a new instance of stroke at the same time or after their injury?
  - 0 - No
  - 1 - Yes
- SWLSCondF: Satisfaction with Life Scale (SWLS) - The conditions of my life are excellent:
  - 1 - Strongly Disagree
  - 2 - Disagree
  - 3 - Slightly Disagree
  - 4 - Neither Agree nor Disagree
  - 5 - Slightly Agree
  - 6 - Agree
  - 7 - Strongly Agree
- SWLSIdealF: Satisfaction with Life Scale (SWLS) - In most ways my life is close to my ideal:
  - 1 - Strongly Disagree
  - 2 - Disagree
  - 3 - Slightly Disagree
  - 4 - Neither Agree nor Disagree
  - 5 - Slightly Agree
  - 6 - Agree
  - 7 - Strongly Agree
- SWLSImprtF: Satisfaction with Life Scale (SWLS) - So far I have gotten the important things I want in life:
  - 1 - Strongly Disagree
  - 2 - Disagree
  - 3 - Slightly Disagree
  - 4 - Neither Agree nor Disagree
  - 5 - Slightly Agree
  - 6 - Agree
  - 7 - Strongly Agree
- SWLSSAtF: Satisfaction with Life Scale (SWLS) - I am satisfied with my life:
  - 1 - Strongly Disagree
  - 2 - Disagree
  - 3 - Slightly Disagree
  - 4 - Neither Agree nor Disagree
  - 5 - Slightly Agree
  - 6 - Agree
  - 7 - Strongly Agree
- SWLSTOT4F: Satisfaction with Life Scale (SWLS) - Satisfaction with life total score using 4 items
  - #
- SWLSTOTF Satisfaction with Life Scale (SWLS) - Satisfaction with Life Scale Total Score
  - #
- tbiInjuryF: OSU TBI Identification Method-Short Form (OSU TBI-ID-SF - In your lifetime, have you ever had any head or neck injury? [Do NOT include the index injury]
  - 0 - No
  - 1 - Yes
- WordRecallCorrectF: Brief Test of Adult Cognition by Telephone (BTACT) - Word Recall: Word recall total correct
  - #
- WordRecallCorrectF_i_n: Brief Test of Adult Cognition by Telephone (BTACT) - Word Recall: Word recall total correct: standardized by age, sex, and education
  - #
- WordRecallIntF: Brief Test of Adult Cognition by Telephone (BTACT) - Word Recall: Word recall number of intrusions
  - #
- WordRecallMiddleF: Brief Test of Adult Cognition by Telephone (BTACT) - Word Recall: Word recall middle correct
  - #
- WordRecallPrimacyF: Brief Test of Adult Cognition by Telephone (BTACT) - Word Recall: Word recall primacy correct
  - #
- WordRecallRecencyF: Brief Test of Adult Cognition by Telephone (BTACT) - Word Recall: Word recall recency correct
  - #
- WordRecallRepF: Brief Test of Adult Cognition by Telephone (BTACT) - Word Recall: Word recall number of repetitions
  - #
